# SARS-CoV-2 genomic and subgenomic RNAs in diagnostic samples are not an indicator of active replication

**DOI:** 10.1101/2020.06.01.20119750

**Authors:** Soren Alexandersen, Anthony Chamings, Tarka Raj Bhatta

**Affiliations:** Geelong Centre for Emerging Infectious Diseases, Geelong, VIC 3220, Australia; Deakin University, Geelong, VIC 3220, Australia; Barwon Health, University Hospital Geelong, Geelong, VIC 3220 Australia

## Abstract

Severe acute respiratory syndrome coronavirus-2 (SARS-CoV-2) emerged in China in late December 2019 and has spread worldwide. Coronaviruses are enveloped, positive sense, single-stranded RNA viruses and employ a complicated pattern of virus genome length RNA replication as well as transcription of genome length and leader containing subgenomic RNAs. Although not fully understood, both replication and transcription are thought to take place in so-called double-membrane vesicles in the cytoplasm of infected cells. We here describe detection of SARS-CoV-2 subgenomic RNAs in diagnostic samples up to 17 days after initial detection of infection, provide evidence for their nuclease resistance and likely protection by cellular membranes consistent with being part of virus-induced replication organelles. Furthermore, we show that the ratios of genomic to subgenomic RNA as well as the ratios of plus to negative strand RNA of genomic and subgenomic RNA are consistent with what have been detected for other coronaviruses in cell culture; albeit with the caveat that *in vivo* diagnostic samples, even in relatively early infection, the ratios of these RNAs are most reminiscent of late culture, semi-purified virus preparations shown to have a relatively constant ratio of genomic to subgenomic RNAs of around 5-10 or higher, while the ratios of positive to negative strands are more than 100 for the genomic RNA and around 20 for the subgenomic RNAs. Overall, our results may help explain the extended PCR positivity of some samples, and may also, at least in part, help explain discrepancies in results of different diagnostic PCR methods described by others; in particular for samples with a low virus load or of poor quality. Overall, we present evidence that subgenomic RNAs may not be an indicator of active coronavirus replication/infection, but that these RNAs, similar to the virus genome RNA, may be rather stable, and thus detectable for an extended period, most likely due to their close association with cellular membranes.

## Introduction

Human coronavirus disease 2019 (COVID-19) emerged in late December 2019 in Wuhan, Hubei Province, China ^1,2^ and a novel betacoronavirus, subsequently named severe acute respiratory syndrome coronavirus-2 (SARS-CoV-2), shown to be the cause. This virus could rather easily transmit from person to person and rapidly spread worldwide ^3,4^. SARS-CoV-2 belongs to the Order *Nidovirales*, Family *Coronaviridae*, Subfamily *Orthocoronavirinae*, Genus *Betacoronavirus*, Subgenus *Sarbecovirus*, Species *Severe acute respiratory syndrome-related coronavirus* and Individuum SARS-CoV-2 with the addition of the strain/sequence, e.g. SARS-CoV-2 Wuhan-Hu-1 as the reference strain ^5^.

Similar to other coronaviruses, SARS-CoV-2 is an enveloped, positive sense, single stranded RNA virus with a genome of nearly 30,000 nucleotides ^6^. After having entered the host cell, replication of coronaviruses initially involves generation of a complementary negative sense genome length RNA for amplification of plus strand virus genome RNA as well as transcription of a series of plus strand subgenomic RNAs all with a common leader joined to gene sequences in the 3’-end of the virus genome. Virus replication and transcription both involve cytoplasmic membrane structures facilitated by virus replication/transcription complexes including virus proteins encoded in the 5’ two thirds of the virus genome (termed Open Reading Frame (Orf) 1a and 1b) by a minus 1 ribosomal frameshift between Orf1a and 1b, and proteolytic processing of the generated polyproteins, and translated from the full length plus sense virus genome RNA. A set of subgenomic RNAs are also generated, most likely from a complex mechanism involving paused negative sense RNA synthesis leading to a nested set of negative sense RNAs from the 3’end of the virus genome joined to a common 5’-leader sequence of approximately 70 nucleotides ^7^. These nested negative sense RNAs in turn serve as templates for transcription of plus strands thus able to serve as a nested set of virus mRNAs for translation of specific proteins from the 3’-third of the virus genome ^7^. These subgenomic mRNAs of SARS-CoV-2, as illustrated in Kim et al. ^8^, are thought to encode the following virus proteins: structural proteins spike (S), envelope (E), membrane (M) and nucleocapsid protein (N) and several accessory proteins for SARS-CoV-2 thought to include 3a, 6, 7a, 7b, 8 and 10 ^8^. The mechanism for generation of these subgenomic mRNAs, are not fully understood ^7,9^, but thought to be tightly regulated to ensure the optimal ratio of virus proteins and to involve pausing of the virus replication/transcription complex at so-called transcription-regulatory sequences (TRS) located immediately adjacent to open reading frames for these virus genes ^8,10^. Furthermore, it appears that the expression of the N protein is required for efficient coronavirus subgenomic mRNA transcription ^7^. The subcellular site/s of coronavirus RNA replication and transcription in the cytoplasm of infected cells is not fully defined, but thought to involve so-called “double-membrane vesicles” (DMV) in or on which the virus replication complex synthesise the needed double and single stranded full length genomic and subgenomic RNAs ^7,11,12^. While it is still unclear whether this RNA synthesis takes place inside or on the outside of these vesicles, it is thought that the membranes somehow “protect” the synthesised RNA, including double stranded RNA, from host cell recognition and response, and also from experimental exposure to RNase ^11,13^. In addition, it has been shown that coronavirus cytosolic RNA is protected from so-called “nonsense-mediated decay” (NMD) by the virus N protein and thus are more stable in that environment compared to what would have been expected for non-spliced RNA ^14^.

While it was originally thought that coronavirus virions contained subgenomic RNAs in addition to the virus plus strand genomic length RNA, it has now been shown that these subgenomic RNAs do not contain a packaging signal and are not found in highly purified, cellular membrane free, coronavirus virions ^15^. However, it is important to stress, that unless specific steps to remove cellular membranes are used for sample preparation and virion purification, such subgenomic coronavirus RNAs are tightly associated with membrane structures, and less purified coronavirus preparations are well known to include subgenomic RNAs that, similar to virion RNA, are nuclease resistant ^16^.

The exact mechanism for generation of coronavirus genomic and subgenomic RNAs are not fully understood, but thought to involve the virus replication complex, the TRS, the N protein and double-membrane vesicles in the cytoplasm of infected cells. Although one study has been published looking at the abundance of subgenomic RNAs for SARS-Cov-2, that study employed virus culture in Vero cells ^8^. That study indicated that while the predicted spike (S; Orf2), Orf3a, envelope (E; Orf4), membrane (M; Orf5), Orf6, Orf7a and nucleocapsid protein (N; Orf9) subgenomic RNAs were found at high levels in cell culture, only low levels of the Orf7b subgenomic RNA was detected and the Orf10 subgenomic RNA was detected at extremely low level (1 read detected, corresponding to only 0.000009% of reads analysed) ^8^. This far, little has been published in regards to the presence of SARS-CoV-2 subgenomic RNAs in samples from infected people. As single study by Wölfel et al 17, looked specifically for the presence of the E gene subgenomic RNA by a specific PCR and took the presence of subgenomic RNA as an indication of active virus infection/transcription. That study could detect E gene subgenomic RNA at a level of only 0.4% of virus genome RNA, in sputum samples from day 4-9 of infection, but only up to day 5 in throat swab samples ^17^. However, while that study assumed a correlation between the presence of the subgenomic E mRNA and active virus replication/transcription and thus active infection, this assumption may not be accurate considering what has been mentioned above about the membrane associated nature of coronavirus RNA and their stability/protection from the host cell response and from RNases. We here describe the detection of SARS-CoV-2 subgenomic RNAs in routine diagnostic oropharyngeal/nasopharyngeal swabs up to 17 and 11 days after first detection by next generation sequencing (NGS) and PCR, respectively, and extend the study to also look at subgenomic RNAs being present in selected read archives selected from the NCBI Sequence Read Archive. Our finding of extended detection of subgenomic RNA in diagnostic samples has subsequently been supported by another study (available as preprint) ^18^ using the same E gene PCR mentioned above ^17^. That very recent study detected subgenomic E RNA in swab samples from hospitalised patients up to 22 days after onset of clinical symptoms ^18^. Thus, it is becoming clear that the presence, and thus detection, of SARS-CoV-2 subgenomic RNAs in diagnostic samples is rather prolonged and consequently not a good marker/indication of active virus replication/transcription or active/recent infection. Despite that, a number of high-profile studies ^19-22^ have continued to use presence or reduction of subgenomic RNA level as evidence of or protection from active infection, and consequently, we believe it is important to understand that these subgenomic RNAs may be present for a significant time after active infection.

Consequently, we here describe detailed analysis of NGS results combined with testing using PCR assays to detect and semi-quantitate SARS-CoV-2 genomic and subgenomic RNAs and to investigate the presence and ratio of positive to negative sense virus and subgenomic RNA. We also present evidence to substantiate that these subgenomic RNAs, similarly to virion genomic RNA, are highly protected from nuclease degradation, most likely by cellular membrane structures, i.e. possibly by so-called double-membrane vesicles known to be important for coronavirus replication and transcription. Finally, by combining the overall results, we conclude that diagnostic swab samples analysed for routine diagnostic purposes are more alike partially purified coronavirus virion preparations from late infection cell culture supernatants, as they appear to contain a relatively stable composition and ratio of full length genomic and subgenomic RNAs, with most full length virus RNA being of positive sense while the subgenomic RNAs have only around 20-fold more positive than negative strand RNA; consistent with these being part of previously active virus transcription complexes protected by double-membrane vesicles.

As both genomic and subgenomic RNAs are present and rather stable in routine diagnostic swab samples, this may explain the extended period of PCR positivity observed in infected individuals and also, at least in part and particularly for samples with a low virus load or of poor quality, explain conflicting findings around “reinfection” as well as discrepancies among diagnostic PCRs detecting different targets in the SARS-CoV-2 genome ^23,24^.

## Results

### Detection, classification and abundance of NGS reads mapped to subgenomic RNAs in SARS-CoV-2 positive samples

As indicated in the Materials and Methods, in our analysis of subgenomic RNAs we have included 12 SARS-CoV-2 positive swab samples of which two were amplified using two different polymerases (for a total of 14 positive sample NGS libraries) included in our study together with a virus-negative control sample (Table 1). Manual inspection of reads indicated the presence of subgenomic RNAs and mapping against a reference (Supplementary Information S1 [file: Wuhan-Hu-1-NC_045512-21500-and-subgenomics-SA4.fasta] also available at NCBI Sequence Read Archive (SRA): PRJNA636225) designed to specifically map the 10 potential subgenomic RNAs, indicated the presence of subgenomic RNAs in all SARS-CoV-2 positive samples while no reads were found in the negative control sample. Interestingly, the abundance of different subgenomic RNAs and the overall number of these subgenomic RNAs varied widely among the 14 samples as indicated in more details below and summarised in Table 2 and Figure 1. Overall, of the 56 million NGS reads generated from the 14 virus-positive samples, nearly 800,000 reads mapped to one of the 10 SARS-CoV-2 subgenomic RNAs (Table 2A). However, no reads mapped to the tentative Orf10/15 RNA and only 5 reads were mapped to the tentative Orf7b RNA. In contrast, reads were mapped to the other 8 subgenomic RNAs, and although it differed among samples, S (Spike), Orf3a and M were consistently mapped at a low level followed in increasing order by subgenomic RNAs for Orf8, Orf6 and E while Orf7a and N were mapped in the highest abundance, although this was not consistent for all samples (Table 2B and Figure 1). As indicated above, the abundance, although overall more or less as expected based on assumed subgenomic RNA abundance ^7,8,10,11,16^, differed widely among samples, most likely depending on sample quality and overall virus genomic and subgenomic RNA abundance. However, comparing 2 samples amplified with two different polymerases generated a somewhat similar picture although overall abundance differed (Table 2B; sample GC-11/34 compared with sample GC-11/38 and GC-14/33 compared with GC-14/37) and comparing different samples with high quality reads and high virus coverage did also, although with some variability from sample to sample, generate a somewhat comparable pattern (Table 2B; samples GC-26/66, GC-11/38, GC-24/61, GC-14/37 and GC-23/60). However, looking at sample quality, as determined by overall average read length of a given sample mapped to the full virus reference genome (Table 2), strongly indicated that sample quality/read length influenced levels of subgenomic RNAs detected, likely due to these subgenomic RNA amplicons incidentally being shorter than many of the virus genome amplicons (Supplementary Table S1 and S2; ThermoFisher SARS-CoV-2 Ampliseq panel include 237 virus amplicons ranging from 54-275 nucleotides in size if amplifying virus genomic RNA). To look at this, we analysed the mapping results of two samples already known to be of poor quality, having been suspended in water rather than PBS/transport medium before coming to our laboratory. Although these two samples had a low Ct (high virus load) in the diagnostic PCRs, the NGS generated mostly very short reads (Table 2B; samples GC-25/65 and GC-55/68) and had a different pattern with a very high abundance of subgenomic RNAs dominated by the Orf7a subgenomic amplicon. This is most likely due to this amplicon being short (sequence length between leader sequence forward primer and nearest pool 2 reverse primer of only 85 nucleotides, although most other subgenomic amplicons would also be expected to be short and some genomic amplicons also being short (supplementary Table S1 and S2). Our sample set analysed here also included samples from two individuals sampled 11-17 days apart and representing early and late infection; sample GC-11/34/38 early and sample GC-24/61 (taken 14 days later) as well as sample GC-20/63 (taken 9 days after the first one, but of poor quality) taken in between those samples and sample GC-14/33/37 early and sample GC-23/60 (taken 11 days later) and sample GC-51/62 (taken 17 days after the first one, but of poor quality), see Table 1 and Table 2. As can be seen when comparing those samples, subgenomic RNAs are detected in the late infection samples and may even be preferentially amplified. Although this may possibly indicate a rather long period of virus replication/transcription, we believe it to be more likely due to coronavirus membrane-associated RNAs being partly, albeit not fully, protected from host and environmental degradation and that samples with partly degraded RNA, represented as shorter average read lengths, have some short subgenomic RNA amplicons preferentially amplified (Table 2 and Supplementary Table S1 and S2), and thus more consistent with such samples mainly containing partly degraded virus genomic as well as subgenomic RNAs.

**Table 1.**
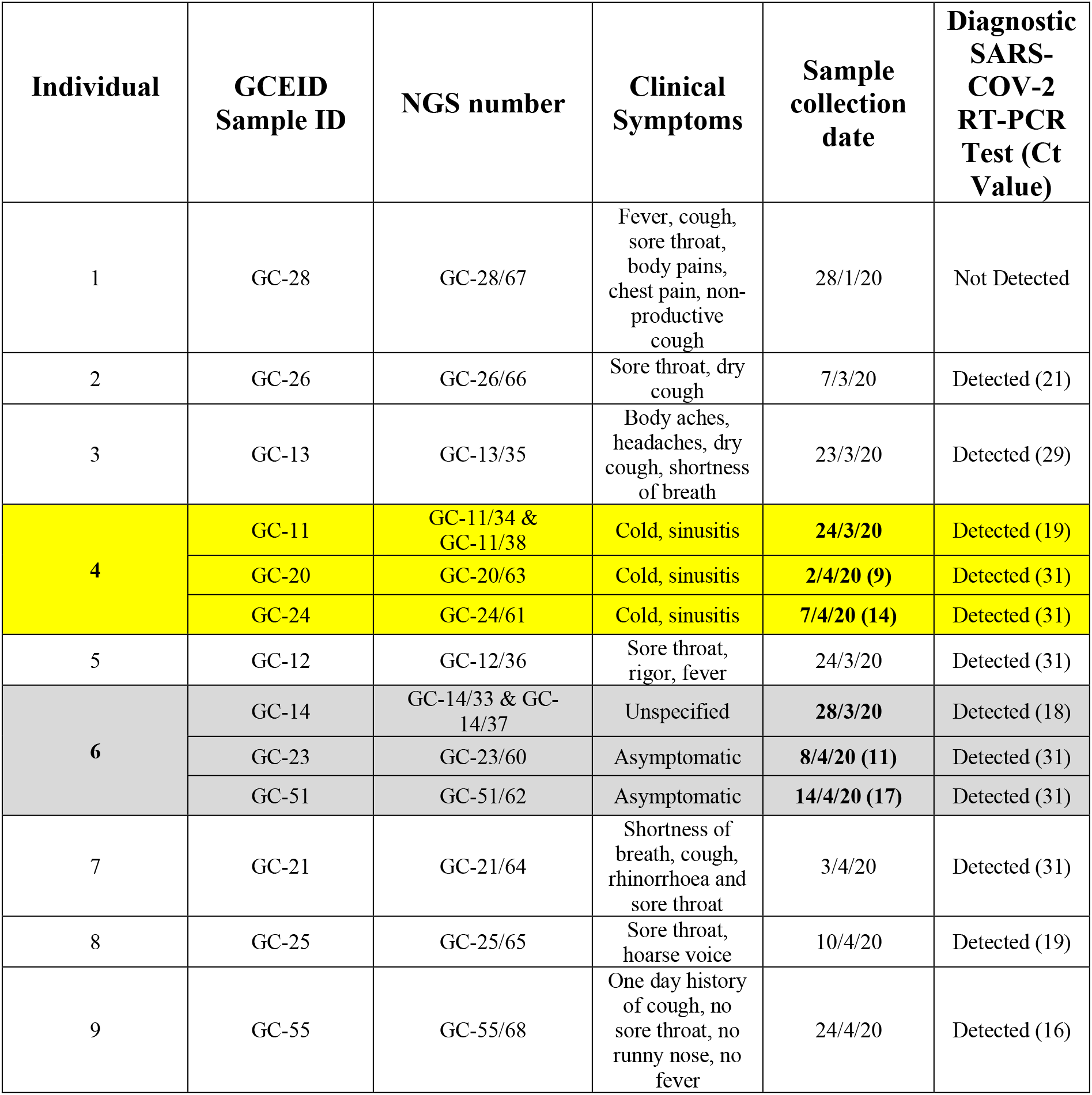
Table showing summary information about the individuals and samples included in this study. Samples from a total of 9 individuals, including one control individual (Individual 1) are included. Two infected individuals (Individual 4 and 6) each had 3 samples collected at different dates and are highlighted in the same colour and in boldface. The days post initial sample collection are shown in brackets after the date. Sample identification and NGS sample number (barcode) is shown together with summary clinical symptoms, sampling date and results of the diagnostic SARS-COV-2 RT-PCR test (Ct value). In part adapted from Bhatta et al. ^28^.

**Table 2.**
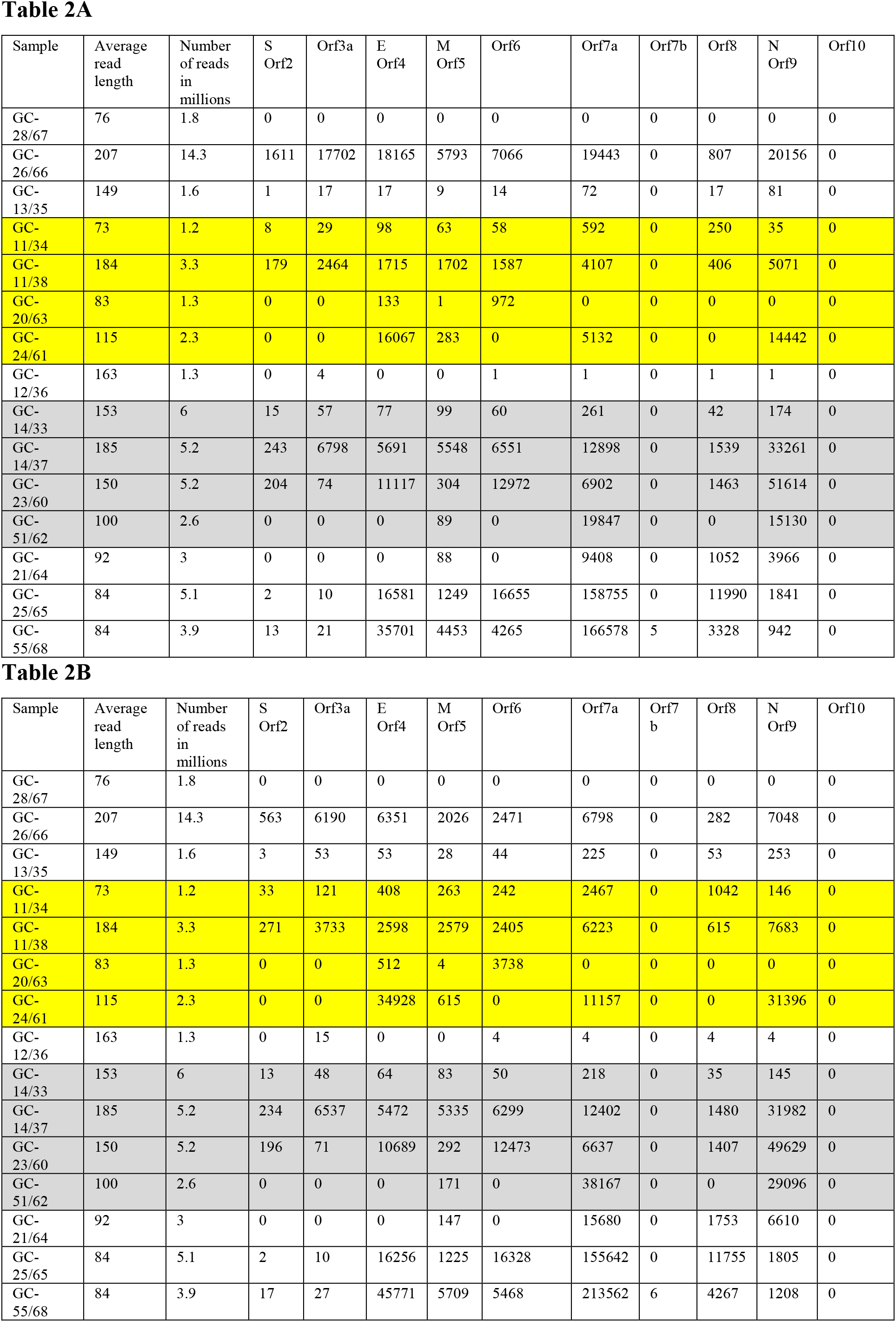
Table showing the details of the sample number, average read length, number of reads and number of reads mapped to each subgenomic SARS-CoV-2 RNA (A). The number of reads given in (B) is adjusted so they represe reads normalised to a total of 5 million reads for each sample for easier comparison. Two infected, individuals each had 3 samples collected at different dates and are highlighted in the same colour and two samples were subjected to NGS using two different DNA polymerases (samples GC-11/34 & GC-11/38 and GC-14/33 & GC-14/37, respectively), see Table 1 for more details.

**Figure 1.**
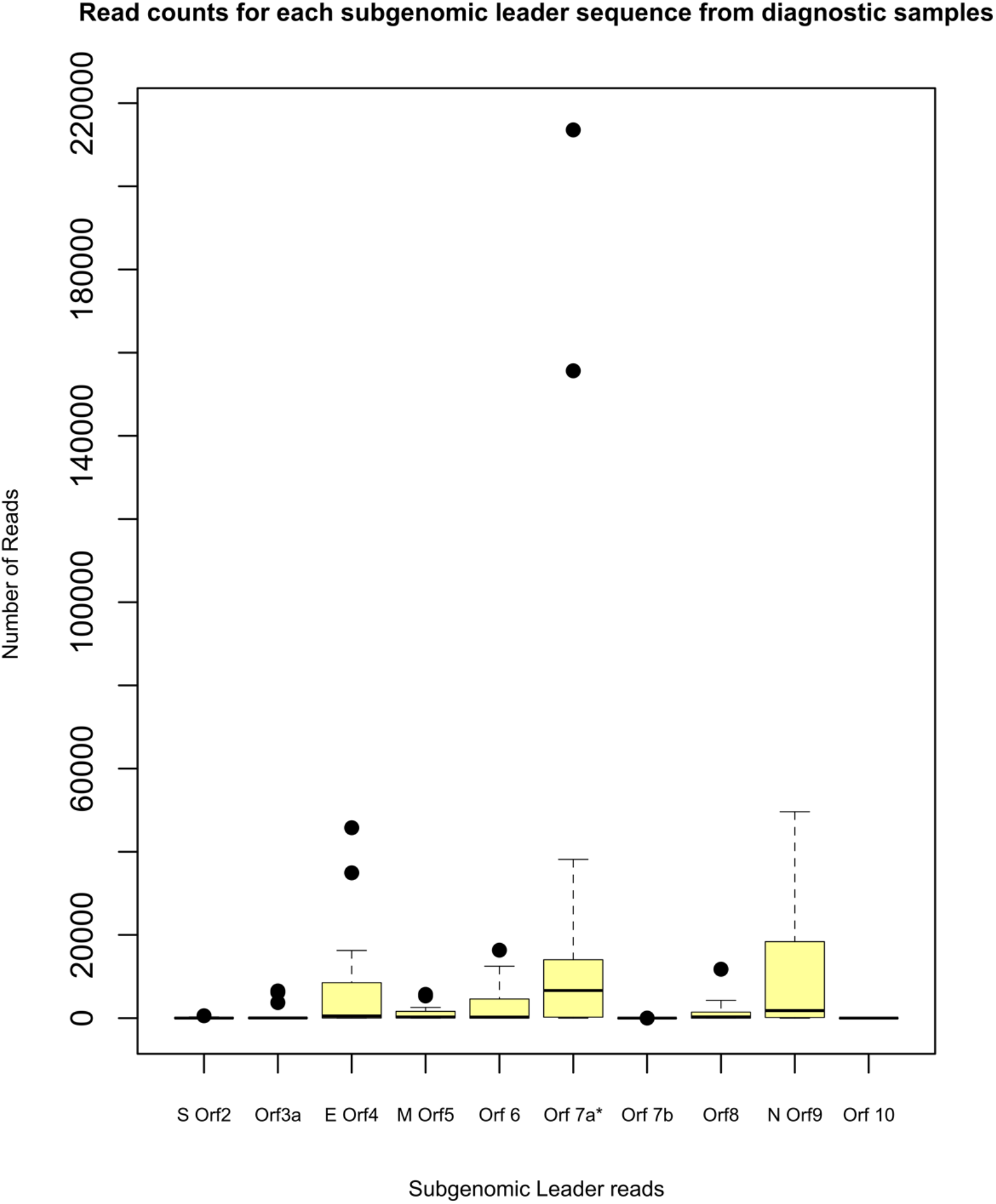
Box-and-whiskers plot showing the number of reads per total of 5 mill reads in diagnostic samples mapped to subgenomic RNAs in the fasta file used for mapping. The median is shown as a line, the box is the 25-75th percentiles and the whiskers are 2.0xIQR (Interquartile Range). Values outside this (outliers) are shown as dark circles.

### Detection of subgenomic RNA reads mapped to the virus genome by filtering reads containing the partial leader sequence

To validate our results detailed above, we looked at the NGS reads to find likely subgenomic RNAs already mapped to the virus reference genome (Wuhan-Hu-1-NC_045512/MN908947.3), but filtering so only reads containing part of the leader sequence were included and then look at where these reads had been mapped. A total of between 8 and 256,123 reads containing the leader sequence were found in our positive samples while none was detected in the negative sample GC-28/67 (Supplementary Table S3). Reads were mapped to the location of the TRS of the 10 known subgenomic RNAs, however, only samples GC-26/66, GC-11/38 and GC-14/37 possessed reads, in a low number, mapping to the start of Orf7b, and only sample GC-26/66 contained any reads mapping to the start of Orf10 (16 reads or 0.001% of total reads in this sample). The number of reads with a leader sequence mapped to the corresponding ORF in the SARS-CoV-2 genome are shown in Supplementary Table S3 and Supplementary Fig. S1. In all samples except samples GC-26/66 and GC-21/64, over 50% of the reads with a leader mapped to the start of a known subgenomic RNA. In samples GC-20/63, GC-24/61, GC-14/37, GC-23/60, GC-51/62, GC-25/65 and GC-55/68, over 75% of reads containing the leader sequence were mapped to subgenomic RNAs (Supplementary Table S3). While, the percentages varied among the samples, the three subgenomic RNAs with the highest median number of reads with the leader sequence were the E gene/Orf4 (4.1%), Orf7a (17.4%), Orf8 (4.3%) and N gene/Orf9 (10.7%).

The samples with the highest number of reads mapping to cryptic or unknown TRS were the poorer quality samples GC-11/34, GC-21/64 and GC-25/65 and no consistent pattern was observed in the mapping of reads with the leader sequence to any individual unrecognised TRS site.

### Searching the NCBI SRA to identify reads mapping to SARS-CoV-2 subgenomic RNAs

Another step in our analysis included searching the NCBI SRA and selection of a few deposited NGS reads from studies using either the same SARS-CoV-2 Ampliseq panel used by us as well as a few generated by different methods, such as direct RNAseq or even virus RNA targeted capture (virus RNA targeted capture SRX8336076; Ion Torrent RNAseq e.g. SRX8346911 USA Vero cells; Ion Torrent Ampliseq e.g. SRX8340472 India; SRX8155154 Tiger USA; MinIon eg ERX4009132 Spain; GridIon SRX788484 SARS-CoV-2/Australia/VIC01/2020, RNAseq oligo-dT selection; Illumina e.g. SRX7777164 Vero cells and SRX7777160 swab USA random PCR). For these selected SRA studies, although not abundant for all of them, reads representing subgenomic RNAs rather than virus genomic RNA could be found by simple analysis using e.g. BlastN. Again, as in our own data, we detected no or very little subgenomic RNA of Orf7b and no evidence for Orf10/15 subgenomic RNA.

To look at this in more detail, we downloaded a selection of SRA’s generated from different sample types, on different sequencing platforms (Illumina, Ion Torrent and Nanopore) and employing different library amplification strategies (Amplicon based, RNA-Seq and random sequencing). Reads covering most of the SARS-CoV-2 genome were present in all archives, except for SRR11454612 which had low coverage over the SARS-CoV-2 genome when mapped to reference MN908947.3. Reads belonging to subgenomic RNA could be identified in all samples except that sample (SRR11454612), representing RNAseq on a sputum sample from an infected human (Supplementary Table S4). The two selected Ion Torrent Ampliseq SRA’s (SRR11810731 and SRR11810737) produced the highest number of subgenomic reads, followed by an RNA-Seq experiment performed in cell culture using a Nanopore platform (SRR11267570). The selected RNA-Seq experiments performed on clinical samples, typically generated very low levels of reads mapping to the virus genome and consequently to the leader sequence. The Artic network primers also detected subgenomic reads in virus culture experiments (ERR4157962 and ERR4157960).

The subgenomic RNAs with the highest median number of reads mapped in the SRA’s were the N gene (10.6%) and Orf7a (7.1%), followed by Orf3a (1.7%) and M gene (1.2%). The subgenomic S gene and Orf6 were typically low (0.1% and 0.9% respectively). No reads were mapped to the subgenomic Orf10 in any sample, and only sample SRR11267570 and SRR11810737 had any reads mapped to the subgenomic Orf7b (0.2-0.3% of reads having the leader sequence).

### Further abundance analysis of SARS-CoV-2 amplicons and cellular gene control amplicons included in the Ampliseq panel

The number of reads in each sample mapped to the individual 237 SARS-CoV-2 genomic RNA amplicons and the included 5 control cellular gene amplicons are shown in Supplementary Table S2. This Table shows the number of reads mapped to each individual amplicon and provides additional information such as the total sum of SARS-CoV-2 reference virus reads, total number of reads mapping to the first 21500 nucleotides of the virus reference genome (thus not overlapping with any potential subgenomic SARS-CoV-2 RNAs) as well as the average number of reads and minimum and maximum number of reads mapped to the individual virus amplicons. Out of the 56 mill total NGS reads from the SARS-CoV-2 positive samples, the total number of reads mapped to the virus amplicons from these samples combined was 31.2 mill reads or 55.7% of all reads of which 16.4 mill reads (29.3% of all reads and 52.6% of reads mapped to the full virus reference genome) mapped to the first 21500 nt of the reference virus genome. For the cellular control amplicons, and in this case including the negative control sample GC-28/67 and thus a total of 58 mill NGS reads, a total of 5.4 mill reads were mapped to these cellular control RNA amplicons corresponding to 9.3% of all reads. Specific details about the abundance of cellular mRNA amplicons, the TATA-box binding protein (TBP NM_003194), LDL receptor related protein 1 (LRP1 NM_002332), hydroxymethylbilane synthase (HMBS NM_000190), MYC proto-oncogene (MYC NM_002467) and integrin subunit beta 7 (ITGB7 NM_000889) in each NGS sample are shown in Table 3. From this analysis it is evident that some samples have nearly no reads mapped to these cellular mRNA amplicons, e.g. samples GC-25/65 and GC-55/68 having been submitted in water, while other samples, such as the low virus load samples GC-23/60, GC-24/61, GC-51/62, GC-20/63 and GC-21/64 and the negative control sample GC-28/67, have many reads mapped to these cellular mRNA amplicons with around half or more of all reads in these samples being mapped (Table 3). Interestingly, samples GC-14/33/37 and GC-11/34/38 also had a low number of reads mapped to cellular mRNA amplicons, however, these samples have a high SARS-CoV-2 load and were taken early in infection. This may also be the case for sample GC-26/66 (Table 3), consistent with a likely reduced level of cellular mRNAs in early, high virus load infection. Sample GC-13/35, and to some extent sample GC-12/36, also had a low number of cellular mRNA amplicon reads which may not be easy to explain as these samples had a relatively low virus load. However, as we only have a single swab sample from these individuals we do not know whether the samples were taken early or late in the infection and moreover, they may simply represent swabs collected without much cellular material.

**Table 3.** Table showing the number of NGS reads, per 5 mill reads, mapped to control cellular mRNA amplicons and the highest and average number of reads for SARS-CoV-2 amplicons included in the Ampliseq panel

| Sample | Sample collection date | SARS-COV-2 PCR (Ct) | TBP | LRP1 | HMBS | MYC | ITGB7 | Total | Mill reads | Total per 5 mill reads | Virus amplicon coverage per 5 mill reads |  |  |
| --- | --- | --- | --- | --- | --- | --- | --- | --- | --- | --- | --- | --- | --- |
|  |  |  |  |  |  |  |  |  |  |  | Highest in first 21500 nt | Highest in full virus genome | Average of 168 amplicons in first 21500 nt |
| GC-28/67 | 28/1/20 | Not Detected | 157551 | 2944 | 70221 | 22633 | 16 | 253365 | 1.8 | 703792 | 0 | 0 | 0 |
| GC-26/66 | 7/3/20 | Detected (21) | 1486 | 193 | 476 | 338 | 732 | 3225 | 14.3 | 1128 | 99927 | 116186 | 48581 |
| GC13/35 | 23/3/20 | Detected (29) | 54 | 2 | 22 | 12 | 120 | 210 | 1.6 | 656 | 3022 | 26200 | 185 |
| GC-11/34 | 24/3/20 | Detected (19) | 0 | 0 | 0 | 1 | 1 | 2 | 1.2 | 8 | 12617 | 3776575 | 726 |
| GC-11/38 | 24/3/20 | Detected (19) | 0 | 9 | 0 | 0 | 2 | 11 | 3.3 | 17 | 55377 | 373552 | 15201 |
| GC-20/63 | 2/4/20 | Detected (31) | 259450 | 11510 | 35621 | 43798 | 284859 | 635238 | 1.3 | 2443223 | 28396 | 1337769 | 72 |
| GC-24/61 | 7/4/20 | Detected (31) | 222864 | 82467 | 66551 | 138145 | 220593 | 730620 | 2.3 | 1588304 | 181672 | 469278 | 9510 |
| GC-12/36 | 24/3/20 | Detected (31) | 421 | 66 | 28 | 626 | 1725 | 2866 | 1.3 | 11023 | 81 | 900 | 4 |
| GC-14/33 | 28/3/20 | Detected (18) | 0 | 1 | 0 | 0 | 1 | 2 | 6 | 2 | 693 | 1880 | 172 |
| GC-14/37 | 28/3/20 | Detected (18) | 1 | 18 | 1 | 2 | 3 | 25 | 5.3 | 24 | 79092 | 91491 | 15358 |
| GC-23/60 | 8/4/20 | Detected (31) | 214567 | 81749 | 91585 | 163657 | 214265 | 765823 | 2.6 | 1472737 | 408398 | 661887 | 21965 |
| GC-51/62 | 14/4/20 | Detected (31) | 483313 | 260623 | 210787 | 159479 | 242317 | 1356519 | 2.6 | 2608690 | 32142 | 269108 | 222 |
| GC-21/64 | 3/4/20 | Detected (31) | 575261 | 167868 | 230305 | 96124 | 580411 | 1649969 | 3 | 2749948 | 125907 | 704475 | 827 |
| GC-25/65 | 10/4/20 | Detected (19) | 5 | 7 | 85 | 186 | 1055 | 1338 | 5.1 | 1312 | 351893 | 1129815 | 3614 |
| GC-55/68 | 24/4/20 | Detected (16) | 2 | 0 | 0 | 0 | 0 | 2 | 3.9 | 3 | 417778 | 946012 | 3922 |

We then looked further at the range of reads mapped to SARS-CoV-2 amplicons in the samples, as the abundance of different size amplicons could possibly be influenced by sample quality or virus load, in particular in poor quality or low virus load samples. The results of this is indicated in the last 3 columns of Table 3 (and further details in Supplementary Table S2). From that data it is evident that certain amplicons are highly abundant, and those amplicons are consistently short amplicons of 68-78 nucleotides (excluding primers) located in the Orf1ab region of the virus genome (the number given in the third last column in Table 3), and thus amplified from genomic RNA, or from one of a few relatively short or from a very short amplicon of 54 nucleotides located in the 3’-end of the virus genome (the number given in the second last column in Table 3), and thus amplifying both genomic and subgenomic RNAs. The average coverage for the 168 amplicons included in the first 21500 nt of the virus genome is shown in the last column of Table 3. In this analysis, sample GC-12/36 stands out as none of the SARS-CoV-2 amplicons are abundant, consistent with this sample having the lowest virus load of the positive samples analysed here (Table 3 and Supplementary Table S2).

### Comparison of abundance of reads mapped to virus and cellular amplicons and abundance of reads mapped specifically to subgenomic RNAs

Due to the variability in individual amplicon abundances observed, we then compared the abundance of reads mapped to SARS-CoV-2 or cellular control amplicons to the abundance of reads mapped specifically to subgenomic RNAs. This comparison was done in a number of different ways as presented below.

One crude comparison, is to compare the around 800,000 reads mapped to the start of subgenomic RNAs to the total of around 31.2 mill reads mapped to the full virus genome or the 16.4 mill reads mapped to the first 21500 nt of the virus genome. This crude comparison indicates that the subgenomic reads with the leader sequence may be around 2.6% of all reads mapped to the full virus genome including from subgenomic RNAs (ratio around 1:40) and 4.9% compared to reads mapped to the first 21500 nt of the virus genome only (ratio around 1:20).

A more realistic comparison of abundance may be to compare the most abundant virus genomic amplicon reads to the most abundant subgenomic reads, or perhaps better to compare the number of reads per amplicon for either genomic or subgenomic reads. First, if using maximum amplicon read for comparison of a likely ratio of genomic to subgenomic RNA reads, i.e. comparing the most abundant amplicon for either purely genomic (first 21500 nucleotides, or for the whole genome (and thus including subgenomic RNAs as well, i.e. both genomic and subgenomics), the ratios are as follows:

Ratio of genomic maximum amplicon (first 21500 nucleotides; Table 3) to most abundant subgenomic amplicon (for six of the samples this is 7a, for 5 samples it is N and for one sample each it is 3A, Orf4 and E; Table 2) is on average 6.0-fold (range 0.8 to 14.2) more genomic than subgenomic RNA. If removing outliers, samples GC-51/62, GC-25/65 and GC-55/68 (low and late sample and the two water samples), the average ratio becomes 7.1-fold more genomic than subgenomic RNA. If doing the same, but using the very most abundant virus amplicon, i.e. whole virus including abundant amplicons close to the 3’-end (Table 3), the ratio of maximum genomic plus subgenomic RNAs to most abundant subgenomic RNA becomes on average 158.5-fold (range 2.9 to 1531). However, removing outliers having a ratio above 100 (samples GC-11/34, GC-13/35 and GC-20/63), then the average becomes 20.6-fold more genomic plus subgenomic RNA than subgenomic RNA only.

In our view, the most meaningful way of comparing abundance of these variable amplicons is to compare the average amplicon abundance for the number of amplicons involved in generating read abundances to be compared. The number of amplicons relevant for the subgenomic RNA reads is 16, i.e. 2 amplicons (1 from each of two pools used for the Ampliseq) for each of the 8 subgenomic RNAs that we are able to detect (for this calculation we do not include orf7b and orf10 as reads for those subgenomic RNAs are essentially zero). However, similar calculations could be made for 20 amplicons and would not change numbers much. The number of amplicons relevant for counting the full virus genome (and including also subgenomic reads as they are also mapped to the reference genome) are 237 and the number of amplicons relevant for only counting genomic RNA, i.e. the first 2/3 of the genome to 21500 nt is 168 amplicons (Supplementary Table S2). Finally, the number of amplicons relevant to count cellular control mRNA amplicons is 10 (although only 5 control amplicons are present in the Ampliseq panel, they are present in both pool 1 and pool 2 and are mapped as one total, i.e. from a total of 10 amplicons).

Based on these numbers of included amplicons, then for the 14 positive samples together having around 800,000 reads mapped to the subgenomic “amplicons” (Table 2), the average per amplicon becomes around 50,000 reads mapped to subgenomic RNA per amplicon for these samples combined.

As we have 31.2 mill reads mapped to the full virus reference genome (and including subgenomic reads) for 237 amplicons that becomes on average 132,000 reads per amplicon while the 16.4 mill reads mapped to the genomic first 2/3 of the reference genome (to 21500 nt) for 168 amplicons becomes an average of around 98,000 reads per amplicon. Finally, the around 5.4 mill reads mapped to the 10 cellular control amplicons becomes an average of 540,000 reads per amplicon. If excluding sample GC-28/67, the SARS-CoV-2 negative control sample and thus not counted in the samples for virus reads above, the control cellular RNA reads becomes slightly less at 515,000 reads per amplicon.

Based on these averages read numbers per amplicon included in the abundance estimates, abundance ratio estimates are as follows:

Virus genomic plus subgenomic to subgenomic RNA only: 2.6

Virus genomic only (i.e. first 2/3^rd^ of the genome to 21500 nt) to subgenomic RNA: 2.0

Control cellular RNA to virus genomic plus subgenomic RNA: 3.9

Control cellular RNA to virus genomic only (to 21500 nt): 5.3

Control cellular RNA to virus subgenomic only: 10.3

Finally, if we look at these ratios and only includes samples GC-14/33/37, GC-11/34/38, GC-13/35, GC-23/60 and GC-26/66, i.e. the samples used for estimation of ratios using PCR (see below and excluding samples GC-25/65 and GC-55/68 that was submitted in water and obviously are outliers in regards to amplicon abundances; Tables 2 and 3 and Figure 1), the ratio of virus genomic plus subgenomic amplicon reads to subgenomic reads only, becomes 6.0 and the ratio of virus genomic amplicon reads only (first 21500 nt) to subgenomic reads only becomes 5.2. If also taking samples GC-14/37 and GC-11/38 out of this assessment, as those samples were subjected to NGS using a different DNA polymerase, these ratios only change slightly to become 6.3 and 5.4, respectively; somewhat similar to what is estimated using PCR, see below.

### SARS-CoV-2 PCR assays to detect subgenomic 7a RNA, genomic and subgenomic 7a RNA and genomic only 5’-UTR RNA

The results for PCR testing of samples, including PCRs for detection of the Orf7a subgenomic RNA including part of the leader sequence; the Orf7a RNA (i.e. covering the 7a open reading frame and consequently detecting any RNA from full length SARS-CoV-2 genomic RNA as well as the subgenomic RNAs of S, Orf3, E, M, Orf6 and Orf7a); the 5’-UTR including part of the leader sequence or only the 5’-UTR downstream of the leader sequence, the latter two assays only detecting SARS-CoV-2 genomic RNA and not subgenomic RNAs, as well as the 3 targets included in the commercial SARS-CoV-2 PCR are shown in Table 4. Of the 12 initial diagnostic positive samples available for testing, 11 were still positive while a single sample previously tested weak positive by PCR and having some SARS-CoV-2 reads by NGS, sample GC-12/36, was now negative, consistent with that sample initially being borderline positive and the cDNA further diluted for this additional PCR testing likely lowering sensitivity with around 1.7-2 Cts (Table 4). Of the 11 PCR positive samples, 7 were positive for all 7 targets tested while another 4 samples were weak positive with high Ct values for only 2-5 out of the 7 targets. These latter 4 samples were all below the detection limit for the 7a subgenomic target while the 7 clearly positive samples were all positive for this target although sample GC-23/60 had a Ct of 35.9 and consequently was only borderline positive (Table 4). Of the 5 samples negative in the 7a subgenomic PCR, this corresponded to the NGS reads for the 7a subgenomic RNA in two of these samples being low or zero (4 and 0 reads per 5 mill NGS reads for samples GC-12/36 and GC-20/63, respectively (Table 2B)). However, the 3 other samples being negative in this PCR (Table 4), samples GC-24/61, GC-51/62 and GC-21/64, had more than 10,000 reads per 5 million NGS reads mapped to the 7a subgenomic RNA by NGS (Table 2B), indicating that the NGS method is more sensitive than PCR for this purpose. This is consistent with these samples only being borderline positive in the other PCRs (Table 4). Interestingly, one sample, sample GC-13/35, that had relatively few 7a subgenomic reads detected by NGS (225 reads per 5 million NGS reads; Table 2B) was weak positive by the 7a subgenomic PCR (Table 4). Overall, the 7a subgenomic PCR was only able to detect the target up to 11 days after first detection while the NGS method also detected a sample taken 17 days after first detection, the last time point included in our study. It should be mentioned though, that we had to dilute the cDNA used for these PCRs as we had limited amounts available, however, for sample GC-13/35, mentioned above and being positive in the 7a PCR, we were able to use undiluted cDNA as we had more cDNA available for that particular sample. Taken together, this indicate that weak positive samples may be close to or just below the threshold of detection by the 7a subgenomic PCR.

**Table 4.** Table showing sample details with corresponding Ct values of PCR amplification using specific targets

| Sample | Sample collection date | Leader-7a Sub-genomic (Set 1) (Ct) | 7a Genomic and Sub-genomic (Set 2) (Ct) | Leader-5'-UTR Genomic (Set 3) (Ct) | 5'-UTR Genomic (Set 4) (Ct) | N Gene (Ct) | Orf 1ab (Ct) | S gene (Ct) |
| --- | --- | --- | --- | --- | --- | --- | --- | --- |
| NTC |  | Neg | Neg | Neg | Neg | Neg | Neg | Neg |
| GC-28/67 | 28/01/2020 | Neg | Neg | Neg | Neg | Neg | Neg | Neg |
| GC-26/66 | 7/03/2020 | 28.9 | 24.8 | 25.7 | 25.9 | 25.7 | 26.2 | 26.7 |
| GC-13/35 | 23/03/2020 | 32.2 | 28.6 | 29.6 | 28.8 | 29.6 | 28.0 | 29.1 |
| GC-11/34/38 | 24/03/2020 | 24.3 | 19.7 | 20.6 | 21.4 | 21.3 | 21.9 | 22.4 |
| GC-20/63 | 2/04/2020 | Neg | 35.3 | 35.6 | 35.7 | 35.8 | Neg | Neg |
| GC-24/61 | 7/04/2020 | Neg | 34.8 | 33.8 | 36 | 35.4 | 35.3 | Neg |
| GC-12/36 | 24/03/2020 | Neg | Neg | Neg | Neg | Neg | Neg | Neg |
| GC-14/33/37 | 28/03/2020 | 21.1 | 17.5 | 17.8 | 18.6 | 19.3 | 19 | 19.7 |
| GC-23/60 | 8/04/2020 | 35.9 | 33.3 | 32.9 | 33.3 | 33.6 | 32.9 | 35.2 |
| GC-51/62 | 14/04/2020 | Neg | Neg | Neg | 35.9 | 37 | Neg | Neg |
| GC-21/64 | 3/04/2020 | Neg | Neg | Neg | 36.8 | 36.3 | Neg | Neg |
| GC-25/65 | 10/04/2020 | 22.3 | 18.9 | 20.1 | 21 | 20.4 | 21.7 | 22.2 |
| GC-55/68 | 24/04/2020 | 19.3 | 15 | 15.7 | 16.1 | 16.4 | 16.7 | 17.5 |

NTC, non-template control (water).

The difference in Ct values between the 7a subgenomic and genomic targets for the 6 samples with a clear positive PCR for both is around 3.4-4.6 Cts, which with the amplification efficiency of these assays corresponds to a difference of around 7-15-fold less of the 7a subgenomic RNA-specific PCR target. However, it should be mentioned, that the so-called 7a genomic PCR, in addition to the virus genomic RNA target, also targets the 7a subgenomic RNA as well as the subgenomic RNAs of S, Orf3, E, M and Orf6. Consequently, these PCRs can only roughly estimate relative abundances. To look at that in more detail, we used two PCRs in the 5’-UTR of the virus designed to only detect virus genomic RNA and not subgenomic RNAs. The results are also shown in Table 4 together with the other PCR results. Comparing the average Ct values obtained by the PCR detecting both the 7a genomic and subgenomic RNAs to those for the two 5’-UTR genomic only PCRs, indicate that subgenomic RNAs may contribute slightly to the PCR signal for the 7a genomic plus subgenomic RNA; however, the difference in Ct values is only 0.44 to 0.99 Ct indicating a ratio of genomic RNA to subgenomic RNAs up to and including 7a of around 1.3-3.3. However, we did not pursue this further as the difference is too small to quantitate with any level of confidence, in particularly considering that the average Ct values for the two 5’-UTR PCRs differed by 0.55 Ct (Table 4), likely indicating that variation within 0.5-1 Ct is to be expected when comparing different PCRs, even if those PCRs are designed to be as similar as possible. In any event, comparing the levels of subgenomic 7a RNA to the levels of genomic plus subgenomic RNAs was not very different from comparing to the genomic (5’-UTR) RNA only (differences within a single Ct). Consequently, for further comparisons we use the results obtained for the two 7a PCRs as they are most comparable and were initially compared when developed using a dilution series of the same amplicon.

## Strand specific PCR

For strand specific PCR, we focused on the 7 samples that were most likely to have a sufficient virus RNA load to allow potential detection of the strand-specificity of the detected SARS-CoV-2 RNA. While all 7 samples were positive for positive sense SARS-CoV-2 RNA, as expected sample GC-23/60 sampled 11 days after first detection (sample GC-14/33/37) was only borderline positive and only for positive sense subgenomic RNA close to the level of detection of these assays (sensitivity approximately 10-fold lower than the non-strand specific PCRs) while sample GC-13/35 was only detected at a relatively low level in the PCR for positive sense genomic RNA (Supplementary Table S5). The identity of positive and negative sense amplicons obtained for sample GC-14/33/37, GC-25/65 and GC-55/68 were confirmed by Sanger sequencing. Only 3 samples were weak positive for negative sense SARS-CoV-2 genomic RNA and of these, only two had a borderline signal for the negative sense 7a subgenomic target. However, it is worth mentioning that these 3 samples included one known to have been taken early in infection, i.e. sample GC-14/33/37, while the other two, also being positive for the negative sense 7a subgenomic target, were samples with a very high virus load according to all diagnostic PCRs and that had been submitted in water rather than in PBS or transport fluid as for the other samples (Samples GC-25/65 and GC-55/68, Supplementary Table S5).

For the samples with a sufficient load for detection by this method, the difference in Ct values for plus strand detection between the 7a subgenomic and genomic targets was around 4.5-5.7 Cts (slightly more than for the non-strand specific PCRs mentioned in the section above (3.4-4.6 Cts), which with the amplification efficiency of these assays correspond to a difference of around 14-28-fold less of the 7a subgenomic positive-sense RNA-specific PCR target. Although this difference in detection of either sense (plus or negative sense) or only positive sense of the subgenomic 7a RNA is small, approximately two-fold, it may possibly indicate that more of the subgenomic RNA as compared to the genomic RNA is of negative sense. Although the number of samples is very low, this is supported by the fact that the difference in Cts obtained between the positive sense and negative sense genomic target is 8.6-8.8 Cts while it is only 5 Cts for the negative sense subgenomic RNA in the two samples for which detectable levels were present ((Samples GC-25/65 and GC-55/68), Supplementary Table S5). As mentioned the sample numbers are low, but may indicate that the ratio of plus to minus sense SARS-CoV-2 genomic RNA is more than 150-fold while that ratio for the 7a subgenomic RNA is only around 20-fold. Furthermore, comparing the Ct values of negative strand genomic to negative strand subgenomic for these two samples, the difference is only 0.9-1.9 Ct, consistent with only around 2-fold more negative strand genomic than subgenomic RNA (Supplementary Table S5). This estimate is based on only two samples, both of which have a very low Ct (high virus target load) in the diagnostic PCRs and both inadvertently having been submitted in water. However, this finding is another indication that subgenomic RNAs may be very stable and not an indication of active replication/transcription, although specific detection of negative strand RNA may be a marker of active/recent replication/transcription if a sufficiently sensitive assay could be established.

To further analyse if samples testing positive for minus sense SARS-CoV-2 RNA potentially contained double stranded RNA, we attempted treatment of samples with the single-stranded RNase If before strand-specific PCRs. The selected samples included samples GC-26/66, GC-11/34/38, GC-14/33/37, GC-25/65 and GC-55/68 and the two strongest samples from the membrane association/fractionation resistance protocol described below. However, after treatment with RNase If, which is described to have a preference for degradation of single-stranded RNA over double-stranded RNA, these samples were completely negative for both positive and negative strand SARS-CoV-2 RNA by PCR. This may be due to a number of factors, including double stranded RNA being below detection limits or possibly that virus plus and negative strands may not have properly annealed before the RNase treatment. Alternatively, the relatively high RNase If concentration used or other nucleases present during incubation of extracted nucleic acids samples at room temperature and at 37 °C in nuclease buffer may have destroyed any double stranded RNA present. We are not able to further look into this as our sample material is now exhausted. However, further studies could look at this in infected cell cultures.

### Membrane association and nuclease resistance of SARS-CoV-2 RNAs

To study a potential membrane association and nuclease resistance of SARS-CoV-2 RNAs, we modified a protocol described for analysis of SARS-CoV replication/transcription complexes in cell culture and applied this protocol to two samples for which we had sufficient volume of sample left and with a sufficient initial SARS-CoV-2 RNA load to allow detection in fractions after applying this protocol. The two samples selected represented two different types of samples, one being sample GC-26/66 representing a good quality sample and the other one being sample GC-55/68, a sample having been suspended in water rather than in PBS or transport medium, and which although having a low Ct (high virus load) as determined by PCR, gave relatively poor SARS-Cov-2 genomic sequence by NGS. It also contained a relatively high number of some subgenomic RNA reads and had almost no reads for the NGS control amplicons representing selected cellular mRNAs.

As part of this protocol, half of the final 16 fractions obtained for each sample were treated with Triton X-100 to determine whether lipid membranes may be important in protecting any SARS-CoV-2 RNA present. As hypothesised, Triton X-100 treatment had a significant effect on final fractions, as the 16 treated fractions became either negative or only borderline positive for all 3 targets in the commercial PCR used, i.e. with Cts in the mid to upper thirties very close to the detection limit of the PCR. Overall, this indicates that the Triton X-100 treatment, even without addition of external nucleases, results in degradation of any SARS-CoV-2 RNA present by at least 1000-fold or more, consistent with such RNA being protected by lipid membranes and consistent with what has been observed for SARS-CoV replication/transcription complexes in cell culture ^25^.

Of the non-Triton X-100 treated fractions, 15 out of 16 had detectable levels of SARS-CoV-2 RNA for all 5 PCR targets used. The only fraction with a single negative result for the 7a-subgenomic PCR was sample GC-26/66 S1S10T-N+ (supernatant from initial 1,000xg spin, non-Triton treated, nuclease treated and then supernatant from final 10,000xg spin). This particular fraction was positive for the other PCR targets, see Supplementary Table S6. Interestingly, but as hypothesised, the good quality sample, sample GC-26/66, ended up with most of the SARS-CoV-2 targets in the final pelleted fractions and these targets being highly resistant to nuclease treatment (Supplementary Table S6). In fact, the nuclease treated fraction (GC-26/66 P1P10T-N+) had a lower Ct, i.e. higher target load than the non-nuclease treated fraction, a phenomenon we have observed earlier for highly purified, nuclease resistant targets ^26^. In contrast, the poor sample, sample GC55/68 that had been in water rather than PBS/transport medium, had most of the targets in the supernatant fraction from the first 1,000xg spin, and what was present in this supernatant was partly susceptible to nuclease treatment and could not be efficiently pelleted by the 10,000xg spin. Furthermore, for this sample the target RNA present in the initial 1,000xg pellet was, in contrast to what was observed for sample GC-26/66, highly susceptible to nuclease treatment (Supplementary Table S6).

Looking at the detected levels of PCR targets for the 7a subgenomic RNA target compared to the 7a genomic target (that detect both genomic and subgenomic targets), the difference was similar to what is indicated in the section above for results directly on samples, around 3-5 Cts, i.e. having around 10 times more genomic than subgenomic targets (Table 4 and Supplementary Table S6). An exception to this pattern may be the initial supernatants subjected to nuclease treatment and then pelleted at 10,000xg (S1P10T-N+). For sample GC-26/66 this fraction had a difference of 5.5 Cts and for sample GC-55/68 of 11 Cts possibly indicating a higher proportion of nuclease protected virion RNA as compared to subgenomic RNAs in this particular fraction (Supplementary Table S6). This would be consistent with the strand specific PCR results for sample GC-55/68 mentioned above, where the Ct difference between positive strand genomic and subgenomic RNA was 8.8 Cts (Supplementary Table S5) and thus also consistent with a high proportion of positive sense virion RNA in that sample.

To look at this in more details, we also did the strand-specific PCR for these fractions. However, the sensitivity of the strand-specific assay (approximately 10-fold less sensitive than the non-strand specific assay) on fractionated samples was not sufficient to detect any negative sense SARS-CoV-2 RNA and no 7a subgenomic RNA could be detected either as it was below the detection limit. However, we could detect positive sense genomic plus subgenomic RNA in most of the fractions in an amount consistent with expected levels detected in the non-strand specific PCRs (Supplementary Table S6, last column).

## Discussion

We here describe the specific detection and mapping of SARS-CoV-2 leader-containing subgenomic RNAs in routine diagnostic oropharyngeal/nasopharyngeal swabs subjected to next generation sequencing (NGS). We present results from two different approaches, one mapping directly to the expected sequences of the leader containing subgenomic RNAs and another approach where reads already mapped to the virus reference genome are filtered based on whether they contain the partial leader sequence or not, to map subgenomic RNAs. We also analyse subgenomic RNAs presence in selected read archives from the NCBI Sequence Read Archive. Furthermore, we extend our study of routine diagnostic samples to include further analysis of NGS reads abundances, semi-quantitation of subgenomic and genomic SARS-CoV-2 RNAs by specific PCRs, quantitation of plus strand as compared to negative strand SARS-CoV-2 RNAs and finally, present results supporting our hypothesis of cellular membrane association and nuclease resistance of the detected SARS-CoV-2 RNAs. Aided by the current understanding of the cell biology of coronavirus infections (please see Introduction for details and specific references), in particular the known association of virus RNAs with, and at least in partial protection by, cellular double-membrane vesicles (DMVs), we present an integrated interpretation of our results based on detailed analysis of relative abundance of the different subgenomic RNAs in samples collected early and late in infection, samples of different quality and in samples subjected to partial cell/membrane lysis in water or by detergent treatment and fractionation and nuclease treatment. Our integrated interpretation of the results overall, is that both virion and subgenomic RNAs are most likely rather stable *in vivo* and that detection of subgenomic RNAs in clinical samples, importantly, do not necessarily signify active virus replication/transcription, but instead is due to such RNAs being part of membrane vesicles, most likely so-called double-membrane vesicles, and thus relatively stable.

Detailed analysis of the NGS mapping showed that some samples, in particular those with a high virus load, had very few reads mapped to the cellular RNA control amplicons included, while other samples, in particular the negative sample and those with a low virus load, had many reads mapped to these cellular RNA amplicons. Based on the observed high variability in reads mapped to individual amplicons, we then looked further at reads mapped to individual SARS-CoV-2 amplicons in the same panel, in particular looking at very short and very abundant amplicons. Indeed, shorter amplicons from genomic or genomic and subgenomic RNAs were highly abundant in all positive samples except one sample with a very low virus load. Comparing the reads mapped to these particular short SARS-CoV-2 amplicons, to the number of reads mapped to amplicons for the SARS-CoV-2 subgenomic RNAs, we found that the ratio of highest genomic to subgenomic RNA reads, focusing on abundant genomic amplicons, varied from around 2-20 depending on the specific comparison made.

The abundance of SARS-CoV-2 subgenomic to genomic RNA by using specific PCRs set up to detect the 7a or 5’-untranslated regions, with one of the PCRs specifically detecting the 7a subgenomic RNA containing the leader sequence and another detecting both 7a genomic RNA and subgenomic RNAs up to and including the 7a subgenomic RNA for the samples with clear positive results, indicated a ratio of genomic and subgenomic 7a RNAs to 7a subgenomic RNA only of around 7-15 fold, consistent with the ratio estimates obtained from the extended analysis of NGS reads mentioned above. Further testing of these samples using so-called strand-specific PCRs, able to detect either the positive or the negative sense of the SARS-CoV-2 RNAs, indicated that for the positive sense RNA, the ratio of the 7a genomic and subgenomic RNA to subgenomic RNA only is around 14-28 fold while the positive to negative sense ratio for the 7a subgenomic RNA is around 20 and around 150-fold or higher for the genomic RNA. Although the presence of both negative and positive sense RNA in some samples indicated that double-stranded forms of these RNAs may be present, the limited sample volumes available and the lower sensitivity of these methods did not allow us to detect that. However, the results obtained for the NGS reads, the non-strand and the strand-specific PCRs are in general agreement with a ratio of genomic to subgenomic RNA of roughly around 10 and, based on the PCR results, with a ratio of plus to negative strand RNA of around 20 for the subgenomic RNA and 150 or higher for the genomic RNA.

The final aspect we wanted to evaluate in this study, was whether the detected SARS-CoV-2 RNAs detected in the diagnostic samples were protected from nucleases, and whether such protection was likely to be facilitated by cellular membranes as hypothesised. The protocol for this part of the study was based on a study of SARS-CoV replication/transcription complexes in cell culture that showed that such membrane complexes could be pelleted by centrifugation at 10,000xg and protected the virus RNA from nucleases unless disrupted by mild detergent treatment^25^. With a slight modification of this protocol to adapt it to diagnostic samples, we were able to show, similar to the original cell culture study of SARS-CoV, that the SARS-CoV-2 RNAs were, at least in part, protected from nucleases, could be pelleted by 10,000xg centrifugation and that detergent treatment, or even to some extent having a sample in water, would greatly reduce the nuclease protection and ability to be pelleted at 10,000xg. Interestingly, even after fractionation, the ratio of genomic plus subgenomic to subgenomic 7a RNA only was still around 10-fold except for a fraction thought to mainly include nuclease protected virion RNA, for which the ratio may be as high as 600-fold more genomic to subgenomic RNA.

In conclusion, the results described here fully support our assessment that SARS-CoV-2 genomic and subgenomic RNAs are present in diagnostic samples even in late infection/after active infection. Subgenomic RNAs, like virion RNA, are rather stable and are likely protected from nucleases by cellular membranes, for the subgenomic RNAs possibly the so-called double-membrane vesicles known to support coronavirus RNA replication and transcription^7,11,25,27^. Detection of subgenomic RNAs in late infection, as described here up to 11 and 17 days by PCR and NGS, respectively, after first detection, the latest time point available to us, although in contrast to the studies by Wölfel et al^17^, is consistent with the recent findings described by van Kampen et al.^18^, which detected the E gene subgenomic RNA by PCR in respiratory swabs up to 22 days after first day of onset of clinical symptoms. The participants in their study likely had more severe disease than the ones included in our study, as their study focused on hospitalised patients, many of which were in intensive care units, while our study subjects only had minor clinical symptoms and all self-isolated at home ^28^. Nevertheless, although their study detected the E gene subgenomic RNA by PCR while we focused on the 7a subgenomic RNA by PCR and all the subgenomic RNAs by NGS, these studies support each other. Although not directly stated in the van Kempen et al. study^18^, but extrapolated by us based on their PCR figures, the SARS-CoV-2 subgenomic E gene RNA is present in such samples at a ratio of genomic to subgenomic RNA of roughly around 10 and this may also be evident by careful inspection of results presented in additional studies published using the same subgenomic E gene PCR ^19-22^. The detection of subgenomic RNA is therefore not direct evidence of active infection, instead its presence is just detected at lower levels than virion genomic RNA resulting in detection for a shorter period of time unless using e.g. highly sensitive NGS.

In conclusion, we believe that the results taken together fits well with what would be expected from a coronavirus infection based on what is known from cell culture studies. The caveat is that samples from even relatively early infection *in vivo*, as assessed by upper respiratory swab samples, are more alike to a late infection cell culture supernatant or partly purified virion preparation and less like what is found for early intracellular coronavirus RNAs in cell culture. Consequently, when looking at what is known for other coronaviruses and cell culture studies, intracellular subgenomic RNAs may dominate over genomic RNA very early on, with 8-70 times more intracellular subgenomic than genomic RNA at 6-8 hours after infection for infectious bronchitis virus, a gammacoronavirus, and bovine coronavirus, a betacoronavirus, and with at least 10 times more plus sense than minus sense RNA ^13,16,27^. In contrast, the same authors found that extracellular and partly purified coronavirus virion preparations from late cell culture infection, while being RNase resistant and susceptible to detergents, have a much higher genomic to subgenomic RNA ratio of 10-30 or higher and at least 100-fold more, positive rather than negative sense RNA^13,16,27^. Consequently, our findings based on NGS, specific PCR assays and fractionation together with nuclease and detergent treatment, is fully consistent with what has been shown from cell culture infection and fractionation of coronavirus replication/transcription complexes in cellular membrane structures, most likely double-membrane vesicles (DMVs). Thus, SARS-CoV-2 RNA in diagnostic swab samples are likely found as a mixture of virion genomic as well as subgenomic RNAs, both protected from nucleases by virus/cellular membranes and at a ratio of around 10-fold more genomic/virion RNA than subgenomic RNA and a plus to minus sense RNA ratio of around 150-fold or more for genomic/virion RNA and around 20 for subgenomic RNA. This stability of subgenomic RNAs together with the variability observed for different amplicons at low target levels, may at least in part help explain variability/discrepancies of PCR results reported for different diagnostic PCR assays detecting targets in different parts of the SARS-CoV-2 genome ^23,24^. For example, our analysis indicated that some subgenomic RNAs may be more abundantly amplified in poor samples, possibly because of partly degraded RNA in such samples and the increased ability of PCR, including most diagnostic PCRs as well as NGS employing various amplification steps, to amplify short targets. We believe that this is particular evident in the Ampliseq and other multi primer-pair sequencing strategies, because partly degraded targets will increase amplification of short amplicons as the “competition” with longer target amplicons is decreased. This notion is supported by our findings in two diagnostic samples that for unrelated reasons had been suspended in water rather than in PBS or transport medium as for our other samples. These particular two samples (sample GC-25/65 and GC-55/68), in which cells and membrane vesicles were almost certainly partly disrupted ^29^, and thus any coronavirus RNAs exposed to the environment and likely to RNases, were dominated by short reads, i.e. short amplicons, and strikingly, with a very high abundance of reads mapped to some subgenomic RNA amplicons, in particular the Orf7a RNA. Clearly, original sample abundance cannot change just because cellular membranes are lysed by the hypotonic treatment, so the observed increase in these reads may almost certainly be caused by preferred amplification of shorter or more efficient amplicons in such samples. Nevertheless, taken together, it shows that these subgenomic RNAs are present in our diagnostic samples and are rather stable.

Our mapping of specific subgenomic RNAs indicated that samples had no or very low levels of subgenomic RNA specific for Orf7b and that a subgenomic RNA specific for Orf10/15 was absent. This is consistent with what has been described for SARS-CoV-2 in cell culture ^8^. A recent publication ^30^ reported conflicting results for diagnostic samples; however, we believe this to be due to misinterpretation of the results of that study, as NGS reads of only 75 nucleotides in length were mapped to the SARS-CoV-2 genome and assigned to subgenomic RNA regions, completely ignoring the fact that only leader containing reads would be specific for any given subgenomic RNA. Consequently, their interpretation of high levels of e.g. Orf10 is most likely incorrect, and their data simply a reflection of a higher coverage of the 3’-end of the genome which is expected as all subgenomic RNAs extend to the 3’-end of the virus genome. Nevertheless, we emphasise that these studies are not comparable, as we believe the study described by Zhang et al. ^30^ is not specifically mapping or filtering leader-containing subgenomic RNAs but simply reports coverage for the different parts of the virus genome.

In conclusion, by combining knowledge of general coronavirus cell biology and replication/transcription with careful mapping of NGS reads to SARS-CoV-2 subgenomic RNAs and by PCR on clinical samples taken at different times of infection and of different quality, we present information that helps understand prolonged and sometimes inconsistent PCR-positivity and may pave the way for development of better diagnostic PCRs and NGS strategies to define active SARS-CoV-2 infection as opposed to extended presence of what most likely represent highly stable virus genomic and subgenomic RNAs present in, and at least in part protected by, cellular membranes, for the subgenomic RNAs most likely so-called double-membrane vesicles (DMVs). Our findings are likely to be relevant also for other coronaviruses and possibly also other viruses in the Order *Nidovirales*. That coronaviruses, and their RNA, may be extremely resistant when part of a membrane matrix is well known, and was demonstrated for example when porcine epidemic diarrhea virus, also a coronavirus, entered and infected pigs in Canada in early 2014 by feed containing spray-dried porcine plasma ^31,32^. Detergent treatment and ultracentrifugation indicated that this coronavirus RNA was initially bound to membranes, but could be pelleted by ultracentrifugation after detergent release and other studies, including our own using yet another coronavirus, the avian infectious bronchitis virus, further support that detergent treatment will release the coronavirus RNA and make it susceptible to nuclease degradation while passing through 0.8, but not 0.45 micron filtres, support the fact that the majority of such coronavirus RNA is membrane bound ^26^. Consequently, we believe that the methods described here to detect and look at relative abundance of SARS-CoV-2 RNAs in clinical samples together with insights in what is known about coronavirus cell biology overall, will have general interest and applicability not only for SARS-CoV-2, but also for other coronaviruses and related viruses.

## Methods

### Samples

We here describe extended analysis of samples already subjected to next generation sequencing (NGS) at the Geelong Centre for Emerging Infectious Diseases (GCEID). The samples and the results of SARS-CoV-2 genomic consensus sequencing have been described previously ^28^. That study included combined nasopharyngeal and oropharyngeal swab samples collected from individuals in the region of Greater Geelong, Victoria, Australia between the 28^th^ of January to the 14^th^ of April 2020. The study included NGS of 11 SARS-CoV-2 PCR-positive samples and 1 negative sample as control. The 11 PCR-positive samples were obtained from 7 individuals as NGS was done on samples taken at three time points from two individuals to monitor their infection ^28^. For the studies described here we added one additional positive sample collected on the 24^th^ of April 2020. This sample was subjected to NGS in exactly the same way as described for the samples mentioned above ^28^ and briefly described in the section below. Summary details of the samples included are shown in Table 1. The study complied with all relevant ethical regulations and has been approved by the Barwon Health Human Research Ethics Committee (Ref HREC 20/56) and participants provided opt-out consent.

In addition to analysis of the NGS reads obtained from the samples mentioned above, we also searched the National Center for Biotechnology Information (NCBI) Sequence Read Archive (SRA) and used selected SRA studies to support the findings from our own samples.

### Nucleic acid extraction, cDNA synthesis and SARS-CoV-2 Ampliseq NGS at GCEID

Nucleic acid extraction and cDNA synthesis was performed as described including heating extracted nucleic acids at 70°C for 5 minutes and rapid cooling on ice before cDNA synthesis using SuperScript™ VILO™ Master Mix (Thermofisher Scientific, Victoria, Australia) as per manufacturers’ instructions and described previously ^28,33^. Prepared cDNA samples were then amplified using the Ion Ampliseq™ Library Kit 2.0 (Thermofisher Scientific, Victoria, Australia) as described earlier ^34,35^ and a commercially available SARS-CoV-2 Ampliseq panel kindly provided by Thermofisher Scientific, Victoria, Australia. In addition, two of the samples (GC-11 and GC-14) that yielded a low virus coverage by this method, were amplified separately (GC-11/38 and GC-14/37) using essentially the same method, but with the Ampliseq Hi Fi mix replaced with Amplitaq Gold 360 Master mix for the amplification step. Amplification was done for either 21, 27 or 35 cycles depending on the estimated virus load in the samples as described previously ^28^ and libraries prepared and run on Ion Torrent 530 chips on an Ion S5 XL genetic sequencer (Thermofisher Scientific) at a concentration of 50pM as per the manufacturer’s protocols and as described previously ^28,33,36^. As described previously ^28^, generated sequence reads were then mapped to a SARS-CoV-2 reference genome (Wuhan-Hu-1-NC_045512/MN908947.3) using the TMAP software included in the Torrent Suite 5.10.1 ^37^, and virus genomic consensus sequences generated using additional Torrent Suite plugins supplied by Thermofisher Scientific, and visualized in Integrative Genomic Viewer ^38^ (IGV 2.6.3) (Broad Institute, Cambridge, MA, USA). Near complete and partial SARS-CoV-2 genomes were aligned using Clustal-W ^39^ in MEGA 7 software ^40^ and near full length sequences submitted to the Global Initiative on Sharing All influenza Database (GISAID) ^41,42^ (https://www.gisaid.org/) as described in our previous study 28.

### Analyses for the detection of SARS-CoV-2 subgenomic mRNAs in the NGS reads

Although the SARS-CoV-2 Ampliseq panel used for the NGS has been designed to generate near full length SARS-CoV-2 genomic sequences, it uses simultaneous amplification of sample cDNA with a total of 242 primer pairs of which 237 primer pairs cover the near full genome of SARS-CoV-2 and an additional 5 amplicons targeting cellular genes in two primer pools ^28^. Close inspection of all primers included in the panel, indicated that two of the forward primers (specifically the first forward primer in each of primer pool 1 and 2, see Thermofisher Scientific for details) have their 3’-end at SARS-CoV-2 (NCBI Accession Wuhan-Hu-1-NC_045512/MN908947.3) ^2^ nucleotide 42 and 52, respectively, and consequently have a perfect match to a sequence included in the SARS-CoV-2 leader sequence with an estimated 27 or 17 nucleotides downstream of these primers also being part of the leader sequence ^7,8,10,17^. Consequently, we concluded that the Ampliseq panel used here would potentially also amplify SARS-CoV-2 subgenomic RNAs by amplification from these two forward primers together with the closest downstream primer included in the same Ampliseq primer pool. Although this was not evident when assembling full virus genome sequences ^28^, a close inspection of reads around expected subgenomic RNA Transcription Regulatory Sites (TRS) ^10^ indicated that a significant number of NGS reads may have been amplified from subgenomic RNAs rather than from virus genomic RNA. To analyse this in more detail, we first assembled an exploratory composite reference for remapping using the Torrent Suite T-map reanalyse function. This initial assembled reference consisted of a composite reference with one sequence containing the first 21500 nucleotides of the SARS-CoV-2 reference genome used for the initial assembly (NCBI Accession (Wuhan-Hu-1-NC_045512/MN908947.3) ^2^ to map reads most likely corresponding to the virus genome while we in addition, assembled 10 tentative subgenomic RNA sequences containing 28 nucleotides from the 3’end of the leader sequence (of which the first 11 nucleotides would be from the forward primer from primer pool 2, if not enzymatically removed by the NGS process) and this leader then followed by the assumed TRS and gene specific sequence for the next 72 nucleotides. Consequently, this reference contained the first 21500 nucleotides of the virus genome (Wuhan-Hu-1-NC_045512/MN908947.3) as well as 10 composite references corresponding to the assumed 5’-end of the 10 potential subgenomic RNAs; S, Orf3a, E, M, Orf6, Orf7a, Orf7b, Orf8, N and Orf10/15 ^7,8,10^. This initial analysis indicated that this was an efficient way of mapping reads corresponding to subgenomic RNAs, and for our final analysis we updated the subgenomic RNA sequences in this composite reference to include the full leader sequence from nucleotide 1-69 and extended the gene specific sequences to ensure that they would include a reverse primer from each primer pool without extending into the next specific gene sequence. This final composite reference used for mapping then included the first 21500 nucleotides of the SARS-CoV-2 genome and the 10 subgenomic RNA specific sequences, each including the leader and gene specific sequences and having a length of 233-364 nucleotides (Supplementary Information S1 [file: Wuhan-Hu-1-NC_045512-21500-and-subgenomics-SA4.fasta] and also available at NCBI Sequence Read Archive (SRA): PRJNA636225). Abundance of mapped reads were determined in IGV at a minimal alignment score of 60 and a mapping quality (MAPQ) of 84.

The next step in the analysis was to look at whether such subgenomic RNAs could be detected by a somewhat different type of analysis based on using reads already mapped to the full virus sequence and then filter these to only look at reads containing part of the leader sequence. This type of analysis should give an unbiased view as to where reads containing the leader may be located on the genome, whether those sites correspond to the assumed position of the genomic leader and proposed TRS of the leader-containing subgenomic RNAs, whether the abundance somewhat correspond to that found by the method mentioned above and finally, whether any additional subgenomic RNAs or cryptic TRS sites may be detected. Reads within the mapped reads BAM files were filtered on whether they had a MAPQ of 32 or higher and contained the partial leader sequence GTAGATCTGTTCTCT, using a custom script written in BASH 4.4.20 and AWK 4.1.4 (GNU project, www.gnu.org) using samtools 1.7 ^43^. This sequence corresponded to nucleotides 52-67 within the SARS-CoV-2 leader sequence in GenBank sequence Wuhan-Hu-1-NC_045512/MN908947.3 ^2^. Reads with this sequence immediately upstream from the mapped region of the read, or within the 5’ end of the mapped region, were retained. The script then generated a spreadsheet giving the nucleotide position of where the leader sequence of each read finished in relation to the reference genome. These reads were then inspected in IGV. To assign the reads to the corresponding subgenomic RNA, the reads were grouped by the nucleotide position at the end of the leader sequence and tallied in Excel. Based on the end position of the leader mapped to the reference genome, each read was assigned to the corresponding subgenomic TRS. Typically, the leader sequence sat within a soft-clipped portion of each read, although depending on the reference sequence, the Ion Torrent TMAP algorithm did occasionally include the start of the leader sequence within the mapped portion of some reads, and at times included spurious insertions or deletions within this section of the mapping in its attempt to map the leader to the reference. Therefore, any read with the leader ending within 10nt of the start of the known subgenomic TRS sequences were assigned to the respective TRS. Some reads did not map to any known TRS, and these were assigned to an “Unknown TRS”.

The next step in our analysis included searching the NCBI SRA from where we selected a few deposited NGS reads from studies using the same SARS-CoV-2 Ampliseq panel used by us and in addition selected a few generated by different methods. SAM files from 15 SRA accessions were downloaded with the NCBI SRAtoolkit sam-dump 2.8.2 and mapped to the Ampliseq SARS-CoV-2 reference MN908947.3 ^2^ using NCBI Magic-BLAST 1.3.0 with a minimum alignment score of 50 and percentage identity of 90% or higher. The script and analysis method to identify reads containing the leader sequence described above was used on the Magic-BLAST mapped SAM files for each of the SRA archives, and the number of reads corresponding to the start of each subgenomic RNA tallied.

### Further abundance analysis of SARS-CoV-2 amplicons and cellular gene control amplicons included in the Ampliseq panel

As mentioned above, the initial goal of the NGS was to assemble the SARS-CoV-2 genome. To look at generated NGS reads in more details, we assessed the abundance of individual amplicons and the mapping data in more details. It should be mentioned, that processing of NGS reads by the Torrent Suite server initially includes trimming of barcode adapters and removal of low-quality and polyclonal reads. A base calling Phred score reflecting the signal quality at each base is then assigned and reads which have poor quality 3’ ends are trimmed by scanning using a 30nt window until the average base calling quality drops to 15. Very short reads still remaining after this step (8 nucleotides or shorter), are then subsequently also removed. Consequently, all read numbers mentioned in this study are reads that have already satisfied these criteria and remained for further analysis/mapping.

We then checked reads mapped to the SARS-CoV-2 Ampliseq panel employed for the NGS that uses simultaneous amplification of sample cDNA with a total of 242 primer pairs of which 237 primer pairs cover the near full genome of SARS-CoV-2 and an additional 5 amplicons targeting cellular genes (see below and Thermofisher for additional details). Checking of mapped reads indicated that they were all mapped uniquely and thus counted only once. Counting/abundance of reads mapped to the individual amplicons were then done using BEDTools ^44^ with a minimum mapping quality (MAPQ) of 20 and requiring the mapped reads to cover more than 90% of an amplicon, and for the amplicons to cover no less than 70% of the read to be included in the count. This ensured reads were not counted more than once, as amplicons targeted partially overlapping regions of the SARS-CoV2 genome, and some of the smaller virus amplicons, were completely overlapped by a larger amplicon. This was done on all samples and in the same way for all the included 237 SARS-CoV-2 amplicons and the 5 control gene amplicons included in the Ampliseq panel. The 5 control gene amplicons span an intron of each of the following cellular genes, and thus amplify mRNAs for TATA-box binding protein (TBP NM_003194), LDL receptor related protein 1 (LRP1 NM_002332), hydroxymethylbilane synthase (HMBS NM_000190), MYC proto-oncogene (MYC NM_002467) and integrin subunit beta 7 (ITGB7 NM_000889). These control cellular gene amplicons are part of the Thermofisher Ampliseq^TM^ panel, and are automatically mapped as part of the SARS-CoV-2 mapping on the Ion Browser as described above.

### SARS-CoV-2 PCR assays to detect subgenomic 7a RNA, genomic and subgenomic 7a RNA and genomic only 5’-UTR RNA

We designed primers for specific detection of the 7a subgenomic RNA by creating a forward primer in the leader sequence and a reverse primer within the 7a sequence itself. A second PCR targeting the Orf7a (i.e. both primers sitting within the 7a open reading frame and consequently detecting any RNA from full length SARS-CoV-2 genomic RNA as well as the subgenomic RNAs of S, Orf3, E, M, Orf6 and Orf7a) was also developed. Two PCRs specifically targeting the 5’-UTR were developed, one including part of the leader sequence and the other targeting the 5’-UTR downstream of the leader sequence. These two assays were specifically designed to only detect SARS-CoV-2 genomic RNA and not subgenomic RNAs. The primers are listed in Table 5.

**Table 5.**
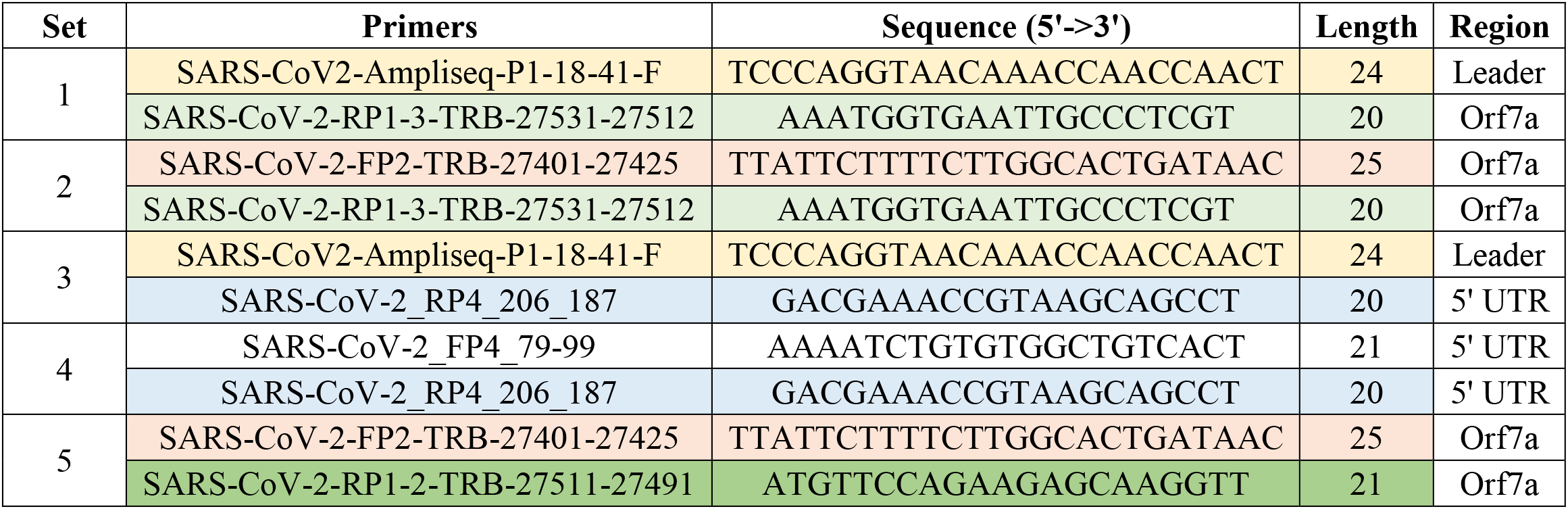
Table showing the primers designed and used to detect specific targets in the SARS-CoV-2 genome.

These PCR assays were all performed using the same cDNA preparations as those used for the NGS; however, as we had limited cDNA volumes remaining for most samples, cDNA was diluted 2.5-fold except for sample GC13/35 for which we had more cDNA available and used undiluted cDNA. The PCRs all employed 2 μl of cDNA and 1X AmpliTaq Gold 360 PCR Mix, 1 μM of each primer, 2 μM Syto 9 (Thermofisher Scientific, Victoria, Australia) and a PCR protocol of 95 °C for 10 min, 40 cycles of 95 °C for 30 sec, 58 °C for 30 sec, 72 °C for 30 sec and a final 72 °C step for 3 min. A melt curve analysis was performed immediately post PCR with the reaction conditions of 95 °C for 15 sec, then 60 °C for 1 min followed by a continuous temperature ramp between 60 °C and 95 °C increasing at 0.05 °C/sec. Positive results were called based on threshold cycle and the correct peak melt temperature of the product. For the initial assay set up, amplicon identity was further confirmed by gel electrophoresis followed by Sanger sequencing of the PCR products as described previously ^33,45^.

In addition to the in-house PCR assays described above, we also used the commercial TaqPath^TM^ COVID-19 RT-PCR Kit (Thermofisher Scientific, Victoria, Australia) using 1.5 μl of the same diluted cDNA samples mentioned above (except for sample GC-13/35 for which we had more cDNA available and used 2.5 μl of undiluted cDNA) and employing the TaqPath^TM^ 1-Step Multiplex Master Mix without ROX (Thermofisher Scientific, Victoria, Australia) together with the TaqPath^TM^ COVID-19 RT-PCR Kit (Thermofisher Scientific, Victoria, Australia) as described previously ^28^ although skipping the initial reverse transcription step. This assay simultaneously detects 3 targets; a target in the Orf1 only detecting the virus genomic RNA, a target in the S gene detecting genomic and S subgenomic RNA and a target in the N gene detecting genomic RNA as well as all full length subgenomic RNAs. Samples were identified as having a high or low virus load based on the Ct obtained from the COVID-19 RT-PCR kit assay or the Ct reported from the original diagnostic laboratory as described previously ^28^.

Efficiency, slope and theoretical sensitivity of each PCR were performed on dilution series of gel purified amplicons for the in-house assays and using a dilution series of a positive control included in the commercial COVID-19 kit. For in house assays using either Syto 9 or SYBR Green (see below), the amplification slopes of the assays were very similar with around 3.9-4.0 cycles between each 10-fold dilution and a lower Ct sensitivity/threshold of 30-32 while the commercial probe-based assay, as anticipated, was more sensitive and efficient with around 3.4 cycles between each 10-fold dilution and a lower Ct sensitivity/threshold of 39 for all 3 targets included.

### Strand specific PCR

For strand specific PCR detection, we used the original nucleic acids extracted for NGS and using an initial step to denature any double-stranded RNA by first heating at 95 °C for 3 min followed by snap-freezing at −20 °C. The samples were then tested using real-time SYBR Green PCR assays with the Power SYBR Green RNA-to-CT 1 step kit (Applied Biosystems, California, USA) using the 7a PCRs described above and adapted so that we initially added a single primer for the reverse transcription step of the protocol at 48 °C for 30 min, then inactivated the reverse transcription enzyme by incubation at 95 °C for 8 min before adding the other primer and continuing the protocol by initially heating to 95 °C for 2 min to further activate the PCR enzyme before conducting 40 cycles of 95 °C for 30 sec, 58 °C for 30 sec, 72 °C for 30 sec and a final 72 °C step for 3 min. A melt curve analysis was performed immediately post PCR with the reaction conditions of 95 °C for 15 sec, then 60 °C for 1 min followed by a continuous temperature ramp between 60 °C and 95 °C increasing at 0.05 °C/sec. Positive results were called based on threshold cycle and the correct peak melt temperature of the product. The initial assay set up was further confirmed by gel electrophoresis of the products followed by Sanger sequencing to confirm amplicon identity essentially as described previously ^33,45^.

To analyse if samples testing positive for minus sense SARS-CoV-2 RNA potentially contained double stranded RNA, we attempted treatment of samples with RNase If (New England Biolabs (NEB), Victoria, Australia) to preferentially remove single stranded RNA before PCR. This was performed essentially as described ^46^, but with a slight modification to promote annealing of extracted RNA before digestion. This was done by adding 1/10^th^ volume of the 10X RNase If buffer (NEBuffer 3) and incubating at room temperature for 10 min before adding 50 units of RNase If and then incubating at 37 °C for 10 min, heating at 95 °C for 3 min followed by snap-freezing at −20 °C to heat inactivate the enzyme and denature any doublestranded RNA before PCR.

### Membrane association and nuclease resistance of SARS-CoV-2 RNAs

To study a potential membrane association and nuclease resistance of SARS-CoV-2 RNAs, we modified a protocol described for analysis of SARS-CoV replication/transcription complexes in cell culture ^25^. This protocol was followed with the following minor modifications. To allow analysis of swab sample material that had already been frozen and thawed at least twice, we started the protocol without an initial Dounce homogenizer step and starting with the swab material in PBS without any additional chemicals or RNase inhibitors. The first step of fractionation consisted of centrifugation at 1,000xg for 5 min and taking the pellet (designated P1, including approximately 10% of the volume of the original sample) and the supernatant fraction (designated S1, approximately 90% of the original volume). The P1 and S1 fractions were then each divided into two aliquots, of which one was treated with 0.5% of the non-ionic detergent Triton X-100 for 15 min at 4 °C. These fractions were then again split into two aliquots of which one was treated with nucleases, first adding a 20 × nuclease buffer and then benzonase and micrococcal nuclease and incubation at room temperature for 30 min as described previously ^26,33^. Fractions were then centrifuged at 10,000xg for 10 min and the pellet fraction (designated the P10 fraction) and the supernatant (designated S10) collected. In effect, this resulted in a total of 16 fractions from each sample of which 8 came from each of the P1 and S1 fractions and of which half had been treated with Triton and the other half not and then half of these fractions treated with nucleases or not. Final fractions were designated P1P10, P1S10, S1P10 and S1S10 and including aliquots that had been treated or not with Triton (T+ or T-) and treated or not with nuclease (N+ or N-). These fractions were then subjected to nucleic acids extraction and cDNA preparation as described for NGS and tested by the 7a subgenomic and genomic PCRs as well as the commercial kit as described above. The obtained PCR values were normalised to the final volume of sample in each of the fractions in order to compare the results. In addition, we also tested the nucleic acids from these fractions in the strand-specific PCRs for 7a genomic and subgenomic RNA.

## Reporting summary

Further information on research design is available in the Nature Research Reporting Summary linked to this article.

## Data availability

The sequence reads and the subgenomic RNA mapping file for our SARS-CoV-2 positive samples reported here have been deposited in the NCBI Sequence Read Archive (SRA) under SRA accession: PRJNA636225. All other data supporting the findings of this manuscript are available in the Supplementary Information files or from the corresponding author upon reasonable request. A reporting summary for this Article is available as a Supplementary Information file.

## Data Availability

The sequence reads and the subgenomic RNA mapping file for our SARS-CoV-2 positive samples reported here have been deposited in the NCBI Sequence Read Archive (SRA) under SRA accession: PRJNA636225. All other data supporting the findings of this manuscript are available in the Supplementary Information files or from the corresponding author upon reasonable request.

## Acknowledgments

We acknowledge clinical and laboratory diagnostic staff for providing samples and for doing the initial diagnostic SARS-CoV-2 testing. We gratefully acknowledge Thermofisher Scientific, Victoria, Australia, for supplying the Ampliseq panel used. We also acknowledge Jason Hodge, laboratory manager of the GCEID laboratory for his technical input. Finally, we gratefully acknowledge the authors and originating and submitting laboratories for the sequence read archives we have used in this study (Supplementary Table S4).

## Author Contributions

S.A. initiated the study, coordinated all work carried out at GCEID and did the initial mapping of SARS-CoV-2 subgenomic RNAs. A.C. did the script-based mapping, mapping of selected SRAs and did the fractionation experiment. T.R.B. provided sample information, participated in the NGS and initial analysis to look for subgenomic RNAs, and did the targeted and strand-specific PCRs. S.A. drafted the initial manuscript with inputs from A.C. and T.R.B. All authors contributed to the final submitted version. All authors have read and agreed to the final version of the manuscript.

## Conflicts of Interest

The authors declare no competing or conflict of interest. The funders had no role in the design of the study; in the collection, analyses, or interpretation of data; in the writing of the manuscript, or in the decision to publish the results.

**Supplementary Materials:** The following is available online; Supplementary Information file S1, Supplementary Tables S1-S6 and Supplementary Figure S1.

## References

1 Hui, D. S. et al. The continuing 2019-nCoV epidemic threat of novel coronaviruses to global health-the latest 2019 novel coronavirus outbreak in Wuhan, China. Int J Infect Dis 91, 264-266 (2020).

2 Wu, F. et al. A new coronavirus associated with human respiratory disease in China. Nature 579, 265-269, doi:10.1038/s41586-020-2008-3 (2020).

3 Chan, J. F.-W. et al. A familial cluster of pneumonia associated with the 2019 novel coronavirus indicating person-to-person transmission: a study of a family cluster. The Lancet 395, 514–523 (2020).

4 Dong, E., Du, H. & Gardner, L. An interactive web-based dashboard to track COVID-19 in real time. The Lancet infectious diseases (2020).

5 Gorbalenya, A. E. et al. The species Severe acute respiratory syndrome-related coronavirus: classifying 2019-nCoV and naming it SARS-CoV-2. Nature Microbiology 5, 536-544, doi:10.1038/s41564-020-0695-z (2020).

6 Wu, F. et al. A new coronavirus associated with human respiratory disease in China. Nature 579, 265–269 (2020).

7 Sola, I., Almazan, F., Zuniga, S. & Enjuanes, L. Continuous and discontinuous RNA synthesis in coronaviruses. Annual review of virology 2, 265–288 (2015).

8 Kim, D. et al. The architecture of SARS-CoV-2 transcriptome. Cell (2020).

9 Snijder, E. J. et al. A unifying structural and functional model of the coronavirus replication organelle: Tracking down RNA synthesis. PLOS Biology 18, e3000715, doi:10.1371/journal.pbio.3000715 (2020).

10 Pascual, M. R. Coronavirus SARS-CoV-2: Analysis of subgenomic mRNA transcription, 3CLpro and PL2pro protease cleavage sites and protein synthesis. *arXiv preprint arXiv:2004.00746* (2020).

11 Snijder, E. J. et al. A unifying structural and functional model of the coronavirus replication organelle: tracking down RNA synthesis. *BioRxiv* (2020).

12 Wolff, G. et al. A molecular pore spans the double membrane of the coronavirus replication organelle. Science, eabd3629, doi:10.1126/science.abd3629 (2020).

13 Hofmann, M. A., Sethna, P. B. & Brian, D. A. Bovine coronavirus mRNA replication continues throughout persistent infection in cell culture. Journal of Virology 64, 4108–4114 (1990).

14 Wada, M., Lokugamage, K. G., Nakagawa, K., Narayanan, K. & Makino, S. Interplay between coronavirus, a cytoplasmic RNA virus, and nonsense-mediated mRNA decay pathway. Proceedings of the National Academy of Sciences of the United States of America 115, E10157-E10166, doi:10.1073/pnas.1811675115 (2018).

15 Escors, D., Izeta, A., Capiscol, C. & Enjuanes, L. Transmissible gastroenteritis coronavirus packaging signal is located at the 5’ end of the virus genome. J Virol 77, 7890-7902, doi:10.1128/jvi.77.14.7890-7902.2003 (2003).

16 Zhao, X., Shaw, K. & Cavanagh, D. Presence of subgenomic mRNAs in virions of coronavirus IBV. Virology 196, 172–178 (1993).

17 Wölfel, R. et al. Virological assessment of hospitalized patients with COVID-2019. Nature, 1-5 (2020).

18 van Kampen, J. J. A. et al. Shedding of infectious virus in hospitalized patients with coronavirus disease-2019 (COVID-19): duration and key determinants. *medRxiv*, 2020.2006.2008.20125310, doi:10.1101/2020.06.08.20125310 (2020).

19 Corbett, K. S. et al. Evaluation of the mRNA-1273 Vaccine against SARS-CoV-2 in Nonhuman Primates. New England Journal of Medicine, doi:10.1056/NEJMoa2024671 (2020).

20 Perera, R. A. P. M. et al. SARS-CoV-2 Virus Culture and Subgenomic RNA for Respiratory Specimens from Patients with Mild Coronavirus Disease. Emerging Infectious Disease journal 26, doi:10.3201/eid2611.203219 (2020).

21 van Doremalen, N. et al. ChAdOx1 nCoV-19 vaccine prevents SARS-CoV-2 pneumonia in rhesus macaques. Nature, doi:10.1038/s41586-020-2608-y (2020).

22 Yu, J. et al. DNA vaccine protection against SARS-CoV-2 in rhesus macaques. Science, eabc6284, doi:10.1126/science.abc6284 (2020).

23 Tom, M. R. & Mina, M. J. To Interpret the SARS-CoV-2 Test, Consider the Cycle Threshold Value. Clinical Infectious Diseases, doi:10.1093/cid/ciaa619 (2020).

24 Xiao, A. T., Tong, Y. X. & Zhang, S. Profile of RT-PCR for SARS-CoV-2: a preliminary study from 56 COVID-19 patients. Clin Infect Dis, doi:10.1093/cid/ciaa460 (2020).

25 Van Hemert, M. J. et al. SARS-coronavirus replication/transcription complexes are membrane-protected and need a host factor for activity in vitro. PLoS Pathog 4, e1000054 (2008).

26 Vibin, J. et al. Metagenomics detection and characterisation of viruses in faecal samples from Australian wild birds. Sci Rep 8, 8686, doi:10.1038/s41598-018-26851-1 (2018).

27 Wu, H.-Y. & Brian, D. A. Subgenomic messenger RNA amplification in coronaviruses. Proceedings of the National Academy of Sciences 107, 12257-12262, doi:10.1073/pnas.1000378107 (2010).

28 Bhatta, T. R. et al. Sequence analysis of travel-related SARS-CoV-2 cases in the Greater Geelong region, Australia. *medRxiv* (2020).

29 Lew, V. L., Hockaday, A., Freeman, C. J. & Bookchin, R. M. Mechanism of spontaneous inside-out vesiculation of red cell membranes. J Cell Biol 106, 1893-1901, doi:10.1083/jcb.106.6.1893 (1988).

30 Zhang, H. et al. Metatranscriptomic Characterization of COVID-19 Identified A Host Transcriptional Classifier Associated With Immune Signaling. Clinical Infectious Diseases, doi:10.1093/cid/ciaa663 (2020).

31 Ojkic, D. et al. The first case of porcine epidemic diarrhea in Canada. Can Vet J 56, 149–152 (2015).

32 Pasick, J. et al. Investigation into the role of potentially contaminated feed as a source of the first-detected outbreaks of porcine epidemic diarrhea in Canada. Transbound Emerg Dis 61, 397-410, doi:10.1111/tbed.12269 (2014).

33 Bhatta, T. R., Chamings, A., Vibin, J. & Alexandersen, S. Detection and characterisation of canine astrovirus, canine parvovirus and canine papillomavirus in puppies using next generation sequencing. Sci. Rep. 9, 1–10 (2019).

34 Chamings, A. et al. Evolutionary analysis of human parechovirus type 3 and clinical outcomes of infection during the 2017–18 Australian epidemic. Scientific reports 9, 1–9 (2019).

35 Alexandersen, S., Nelson, T. M., Hodge, J. & Druce, J. Evolutionary and network analysis of virus sequences from infants infected with an Australian recombinant strain of human parechovirus type 3. Sci. Rep. 7, 1–12 (2017).

36 Bhatta, T. R., Chamings, A., Vibin, J., Klaassen, M. & Alexandersen, S. Detection of a Reassortant H9N2 Avian Influenza Virus with Intercontinental Gene Segments in a Resident Australian Chestnut Teal. Viruses 12, 88 (2020).

37 Caboche, S., Audebert, C., Lemoine, Y. & Hot, D. Comparison of mapping algorithms used in high-throughput sequencing: application to Ion Torrent data. BMC genomics 15, 264 (2014).

38 Thorvaldsdóttir, H., Robinson, J. T. & Mesirov, J. P. Integrative Genomics Viewer (IGV): high-performance genomics data visualization and exploration. Briefings in bioinformatics 14, 178–192 (2013).

39 Larkin, M. A. et al. Clustal W and Clustal X version 2.0. bioinformatics 23, 2947–2948 (2007).

40 Kumar, S., Stecher, G. & Tamura, K. MEGA7: molecular evolutionary genetics analysis version 7.0 for bigger datasets. Molecular biology and evolution 33, 1870–1874 (2016).

41 Elbe, S. & Buckland-Merrett, G. Data, disease and diplomacy: GISAID’s innovative contribution to global health. Global Challenges 1, 33–46 (2017).

42 Shu, Y. & McCauley, J. GISAID: Global initiative on sharing all influenza data–from vision to reality. Eurosurveillance 22 (2017).

43 Li, H. et al. The Sequence Alignment/Map format and SAMtools. Bioinformatics 25, 2078–2079, doi:10.1093/bioinformatics/btp352 (2009).

44 Quinlan, A. R. & Hall, I. M. BEDTools: a flexible suite of utilities for comparing genomic features. Bioinformatics 26, 841-842, doi:10.1093/bioinformatics/btq033 (2010).

45 Chamings, A. et al. Detection and characterisation of coronaviruses in migratory and non-migratory Australian wild birds. Sci. Rep. 8, 1–10 (2018).

46 Wang, P.-H. et al. RNase If-treated quantitative PCR for dsRNA quantitation of RNAi trait in genetically modified crops. BMC Biotechnology 18, 3, doi:10.1186/s12896-018-0413-6 (2018).

